# NMR metabolomics for cardiovascular–kidney–metabolic risk stratification in an integrated healthcare system

**DOI:** 10.64898/2026.10.01.26364322

**Authors:** Jonathan Chacon-Barahona, Carlos Iribarren, Jamal Rana, Gina Rinetti Vargas, Sarah Rowell, Olivier Elemento, Karsten Suhre, Devin Absher, Jan Krumsiek

## Abstract

Metabolomic profiling predicts future disease in population cohorts, but its value in real-world healthcare remains uncertain. We profiled 250 nuclear magnetic resonance (NMR) biomarkers in 54,933 participants from a diverse integrated US healthcare system linked to longitudinal electronic health records, evaluating nine cardiovascular–kidney–metabolic disease endpoints. Adding NMR to standard-of-care predictors improved discrimination for nearly all endpoints, with the largest gains for type 2 diabetes, chronic kidney disease and metabolic dysfunction-associated steatotic liver disease. Beyond disease onset, NMR improved prediction of disease-specific complications and cross-organ outcomes. Gains persisted across demographic, anthropometric, and genetic-risk strata and were greatest where standard-of-care prediction was weakest. NMR-enhanced models generally preserved calibration, improved high-risk enrichment and increased decision-curve net benefit. Together, these findings support NMR metabolomics as a common approach to risk stratification across the cardiovascular–kidney–metabolic disease continuum and illustrate how NMR-enhanced risk estimates could be integrated with longitudinal clinical information.

## 1 Main

Cardiovascular, kidney, and metabolic (CKM) diseases are leading causes of death and disability worldwide, yet identifying individuals at sufficiently high risk to benefit from early intervention efforts remains a major clinical challenge^1–3^. Recent AHA/ACC/ADA/ASN guidance explicitly frames CKM diseases as an interconnected syndrome and emphasizes prevention of progression across this continuum^4^. Risk assessment, however, remains largely disease-specific, relying on combinations of demographic and clinical characteristics, laboratory measurements and prediction tools, including HbA1c for type 2 diabetes, estimated glomerular filtration rate (eGFR) for chronic kidney disease, FIB-4 for liver fibrosis and the American Heart Association PREVENT equations for cardiovascular disease^5–8^. These approaches use a limited set of measurements and show inconsistent discrimination and calibration across populations and healthcare systems^9–11^. Consequently, individuals who later develop disease may not be identified as high risk in time for early evaluation or preventive intervention^12^.

Targeted nuclear magnetic resonance (NMR) metabolomics offers a potential way to address this mismatch by using a single blood test to simultaneously quantify circulating biomarkers across multiple metabolic pathways, at a cost comparable to a routine lipid panel^13^. In UK Biobank, NMR-derived profiles were predictive of incident events for 23 of 24 common diseases and improved discrimination beyond comprehensive clinical predictors for eight of these diseases, with the largest gain for type 2 diabetes and additional gains for heart failure, dementia and COPD^14^. Subsequent work linked NMR biomarkers to more than 700 disease outcomes^15^. Across three national biobanks, NMR-derived scores outperformed polygenic risk scores for many common chronic diseases^16^ and improved cardiovascular risk reclassification when added to European guideline-endorsed equations^17^.

Despite this evidence, metabolomic profiling remains uncommon in routine clinical care^18^. One notable exception is the use of untargeted metabolomics to support diagnosis of inherited metabolic disorders in specialized laboratories^19–21^. More broadly, Finland’s largest private healthcare provider includes an NMR-based blood test in occupational health assessments nationwide^22^. These examples show that metabolomic technologies can be incorporated into clinical workflows but they do not establish whether the predictive value of NMR metabolomics demonstrated in research biobanks translates to routine healthcare, where clinical measurements are irregular and often incomplete^23–25^. Evaluations of common-disease prediction and clinical utility have largely been conducted in population-based research biobanks comprising relatively healthy, predominantly European volunteers, rather than in diverse health-system populations with a substantial burden of chronic disease^26,27^. They have also focused mainly on disease onset, whereas after diagnosis the clinical task shifts to predicting complications and disease progression^16^.

The Kaiser Permanente Research Bank (KPRB) provides a unique opportunity to address this translational gap. Embedded within an integrated US healthcare system, KPRB includes a diverse population with banked serum samples linked to comprehensive longitudinal electronic health records capturing routine laboratory measurements, diagnoses, pharmacy dispensing records, and healthcare utilization^28^. For this study we profiled 250 circulating biomarkers on the Nightingale Health NMR platform in blood samples from 54,933 participants and anchored predictions at the time of blood collection to emulate prospective risk assessment in a realistic clinical setting. Across nine cardiovascular, kidney, and metabolic disease endpoints, we assessed whether NMR-enhanced models improved prediction beyond standard-of-care information, produced predicted risks that matched observed event rates, increased clinical net benefit and performed consistently across demographic, clinical and genetic-risk strata. We further evaluated prediction of complications among patients with established disease to assess risk stratification after diagnosis, and tested whether models developed in UK Biobank generalized to KPRB.

## 2 Results

### 2.1 The Kaiser Permanente Research Bank metabolomics dataset

We profiled serum samples from 54,933 Kaiser Permanente Research Bank (KPRB) participants (Figure 1). The metabolomics cohort was drawn from the broader KPRB population of approximately 440,000 participants^28^, with prioritization to support racial and ethnic diversity, longer longitudinal follow-up and enrichment for cardiometabolic, kidney, and pulmonary disease (Extended Fig. 1).

**Figure 1.**
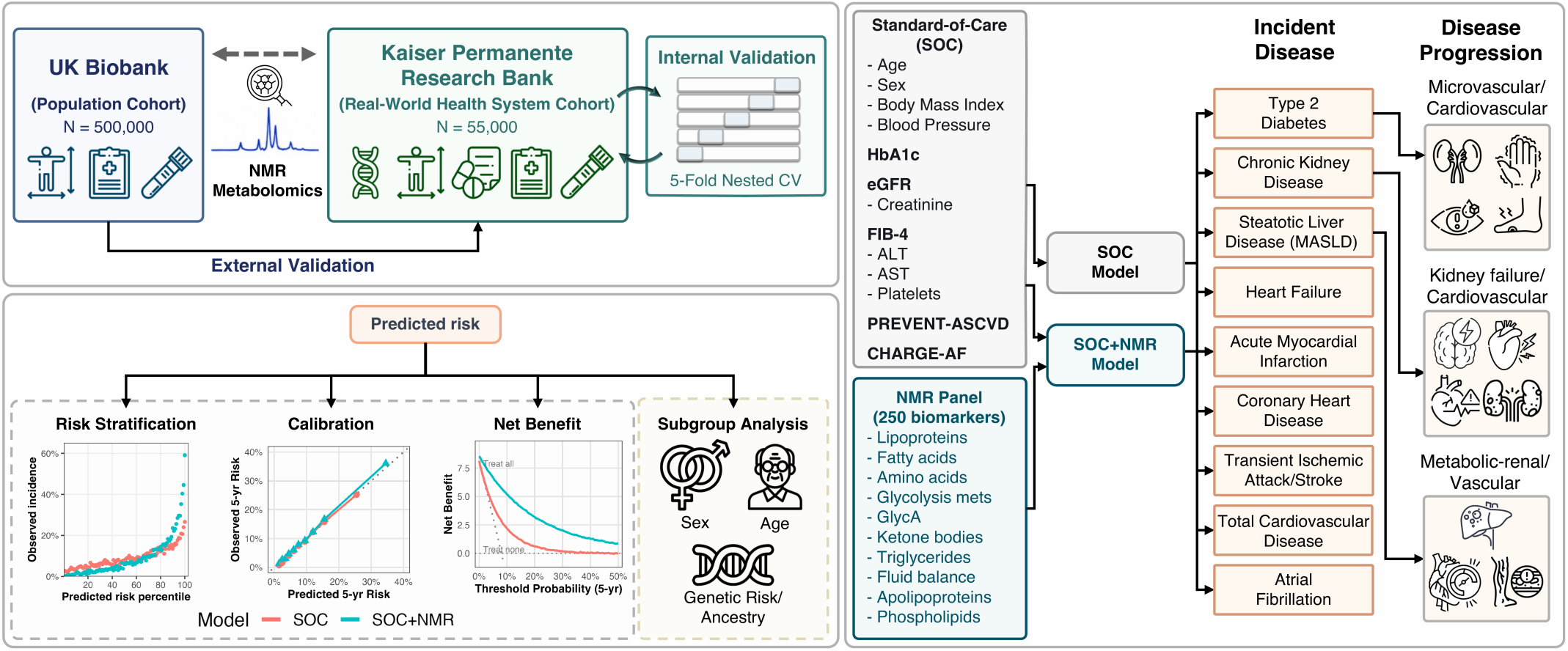
Kaiser Permanente Research Bank NMR metabolomics dataset and analysis framework. Serum ^1^H NMR metabolomics was generated for 54,933 Kaiser Permanente Research Bank participants and linked to longitudinal electronic health records. UK Biobank was used as an independent cohort to assess the transferability of predictive models developed in the Kaiser Permanente Research Bank. The workflow figure summarizes cohort selection, NMR biomarker domains, endpoint ascertainment, and modeling analyses used to evaluate incident disease risk, secondary complications, polygenic risk scores, calibration, and clinical risk prioritization.

At sample collection, participants had a median age of 61.5 years (range 18–89) and were 62% female; by self-reported race and ethnicity, the cohort comprised 62% White, 13% Hispanic, 12% Asian, 9% Black and 4% other racial or ethnic groups (Supplementary Table 1). Median EHR follow-up was 6.8 years, from blood collection to Kaiser membership disenrollment, death or end of follow-up (Supplementary Table 2). Metabolomics data were linked to longitudinal BMI, blood pressure, laboratory measurements, diagnosis codes, medication dispensing, lifestyle survey data and healthcare utilization (Supplementary Table 3).

Metabolomic profiling was performed using the Nightingale Health ^1^H NMR metabolomics platform^14,16,29^, which simultaneously quantifies 250 blood-circulating biomarkers spanning size-resolved lipoprotein triglyceride, cholesterol and phospholipid content, lipoprotein particle numbers, selected fatty and amino acids, glycolysis metabolites, ketones and other small molecules, as well as inflammation-related protein markers (Supplementary Table 4). As part of quality control, metabolite measurements were adjusted for minor signal drift associated with storage duration and sample transport time (Methods; Supplementary Fig. 1). Metabolic markers showed a high degree of correlation (Supplementary Fig. 2), motivating the use of regularized models in subsequent analyses.

### 2.2 Metabolomics improves disease prediction beyond clinical risk factors

We tested whether the use of NMR metabolomic profiles improved prospective risk stratification beyond standard-of-care (SOC) predictors in KPRB. Using the blood sampling timepoint as baseline, we modeled time to first diagnosis across nine CKM endpoints: type 2 diabetes (T2D), chronic kidney disease (CKD), metabolic dysfunction-associated steatotic liver disease (MASLD), heart failure (HF), acute myocardial infarction (Acute MI), coronary heart disease (CHD), transient ischemic attack (TIA)/stroke, Total Cardiovascular Disease (Total CVD), and atrial fibrillation (Afib) (Table 1; Methods).

**Table 1.**
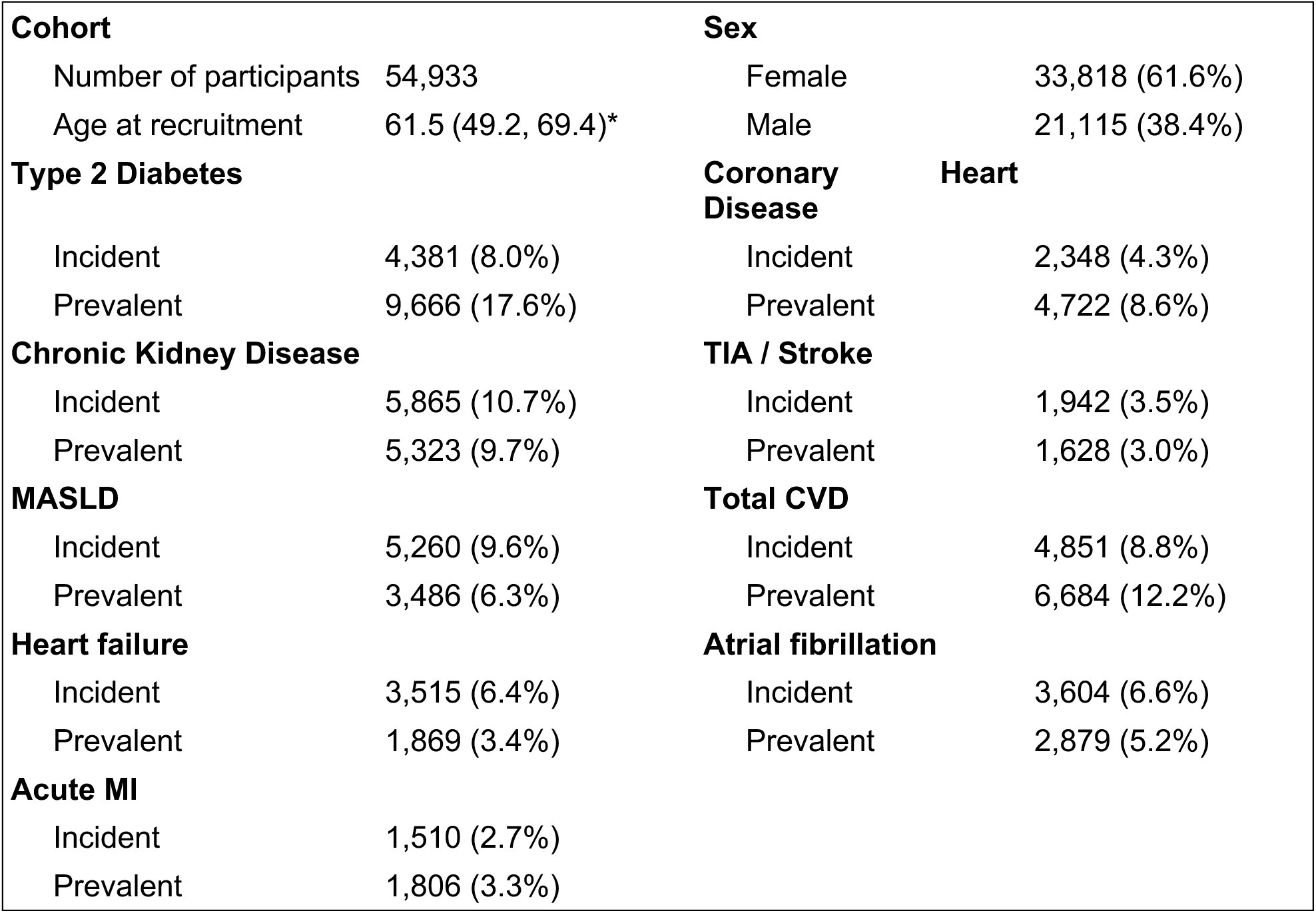
Study characteristics. MI = Myocardial infarction. MASLD = Metabolic dysfunction-associated steatotic liver disease. TIA = Transient ischemic attack. Total CVD = Total Cardiovascular Disease (which includes Coronary Heart Disease, Acute MI, TIA/Stroke, and Heart Failure). Prevalent = Patient was diagnosed with disease before or at time of blood draw. Incident = Patient was diagnosed with disease between time of blood draw and last follow-up in the healthcare system. * Median and IQR (central 50%) shown.

We evaluated NMR across nested predictor sets with increasing clinical information. BASE models included age and sex, available without any clinical testing; CLIN additionally included body mass index and systolic blood pressure, routinely assessed in clinical care; and SOC further included endpoint-specific routine laboratory biomarkers and established clinical risk predictors (HbA1c for T2D, eGFR for CKD, FIB-4 for MALSD, and PREVENT for CVD-related conditions, CHARGE-AF for Afib). All 250 NMR biomarkers were added jointly to each predictor set using ridge-penalized Cox regression. Performance was assessed by five-fold cross-validation using the C-index, which quantifies discrimination between participants with earlier events and those with later or no events.

Across endpoints, adding NMR to SOC models significantly improved incident disease discrimination in nearly all comparisons (Fig. 2a,b). Gains were largest for T2D (ΔC-index +0.07), CKD (+0.08) and MASLD (+0.05), with smaller but consistent improvements for cardiovascular outcomes (+0.005 to +0.02). NMR also improved discrimination for COPD, evaluated as a supplementary pulmonary endpoint (Supplementary Fig 3).

**Figure 2.**
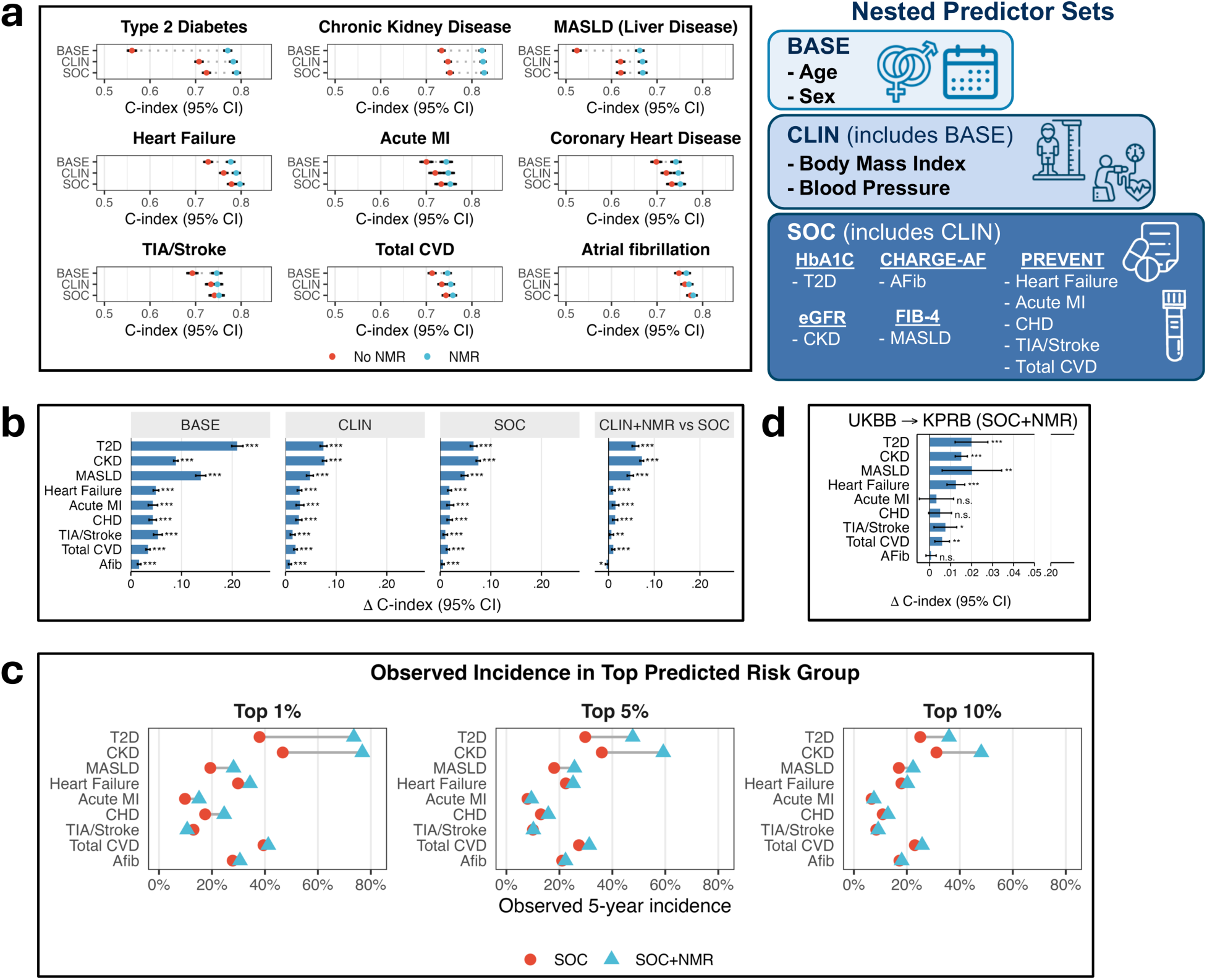
NMR metabolomics improves incident disease risk prediction beyond routine clinical predictors. **a,** C-index estimates and 95% confidence intervals for nested Cox survival models across incident disease endpoints. Predictor tiers were BASE (age and sex), CLIN (BASE plus body mass index and systolic blood pressure), and standard of care (SOC; CLIN plus endpoint-specific routine clinical predictors). NMR-enhanced models added the 250-biomarker NMR panel to each tier. **b,** C-index differences between models with and without NMR across predictor tiers. Also shown is the comparison of CLIN+NMR with SOC, testing whether NMR combined with basic clinical measures outperformed endpoint-specific laboratory-based clinical predictors. **c,** Observed 5-year incidence among participants in the top 1%, 5%, and 10% of predicted risk according to SOC and SOC+NMR models. **d,** C-index differences between models with and without NMR when models were trained in UK Biobank and evaluated in KPRB. In **b** and **d**, error bars indicate 95% confidence intervals. P values were obtained from paired C-index comparisons and adjusted across the nine endpoints within each comparison using the Benjamini–Hochberg procedure. * p<0.05, **p<0.01, ***p<0.001, for FDR-corrected p-values.

Second, we compared CLIN+NMR with SOC models to test whether a single NMR assay could replace endpoint-specific routine laboratory measurements. CLIN+NMR outperformed SOC for eight of nine endpoints, with the largest C-index gains for CKD (+0.07), T2D (+0.06) and MASLD (+0.05), and smaller gains for most cardiovascular and pulmonary outcomes. For atrial fibrillation, CLIN+NMR and SOC models showed nearly identical discrimination (ΔC-index, −0.004) (Fig. 2a,b; CINDEX_STATS_PRIMARY). Detailed regression coefficients and statistics can be found in Supplementary Table 5 and Supplementary Table 6.

Three sensitivity analyses supported the robustness of these findings. Restricting the analysis to participants with complete clinical measurements in the preceding 2 years yielded similar results, suggesting that the observed NMR gains were largely not attributable to the handling of missing data. (Supplementary Fig. 4). Excluding diagnoses within 6 and 12 months modestly reduced absolute discrimination but left NMR improvements largely unchanged, arguing against inflated performance due to occult or near-term disease (Supplementary Fig. 5). Results were also similar after excluding participants without EHR encounters in the preceding 2 years, suggesting that the observed NMR gains were not driven by clinical-data missingness among participants with limited healthcare engagement (Supplementary Fig. 6).

Improved discrimination translated into greater enrichment of incident cases among participants at highest predicted risk (Fig. 2c; Supplementary Fig. 7). In the top 1%, observed 5-year incidence increased from 38.0% to 73.6% for T2D and from 46.7% to 76.8% for CKD with SOC+NMR, with smaller gains for most other endpoints and little improvement for TIA/stroke. Similar enrichment was observed in the top 5% and 10%. NMR-enhanced models also preserved or improved calibration, meaning that predicted event probabilities closely matched observed event rates, across 1-, 3-, and 5-year prediction horizons (Extended Data Fig. 2). For CKD, SOC+NMR underestimated risk at the upper end, indicating that endpoint-specific recalibration may be required before clinical use.

To assess cross-cohort generalizability, we trained the same models in UK Biobank and evaluated them in KPRB after harmonizing serum–plasma differences (Fig. 2d; Supplementary Fig. 8). Although absolute discrimination was lower than for KPRB-trained models, NMR improved discrimination for T2D (ΔC-index +0.020), MASLD (+0.020), and CKD (+0.015), with smaller gains for cardiovascular endpoints and confidence intervals spanning zero for acute myocardial infarction, CHD, and atrial fibrillation. Thus, incremental predictive signal from NMR was retained across cohorts, most clearly for T2D, MASLD, and CKD.

### 2.3 Metabolomics distinguishes risk where standard-of-care prediction is weakest

Having established that NMR metabolomics improved disease prediction across the full KPRB cohort, we next examined model performance within patient subgroups. Population-wide discrimination can appear stronger in heterogeneous populations because the overall C-index reflects both within-group discrimination and comparisons between groups with substantially different baseline risks—for example, older versus younger participants or participants with obesity versus those with lower BMI. Restricting the evaluation to one group at a time reduces this contribution and provides a more stringent, clinically relevant test of whether the models can further distinguish higher- and lower-risk individuals with similar characteristics.

Across endpoints, adding NMR metabolomics improved risk discrimination in nearly every subgroup defined by age, BMI, sex, race-and-ethnicity category, or polygenic risk (Fig. 3; Supplementary Table 7). The largest gains generally occurred in groups in which SOC models had the lowest discrimination, yielding more consistent discrimination across patient groups.

**Figure 3.**
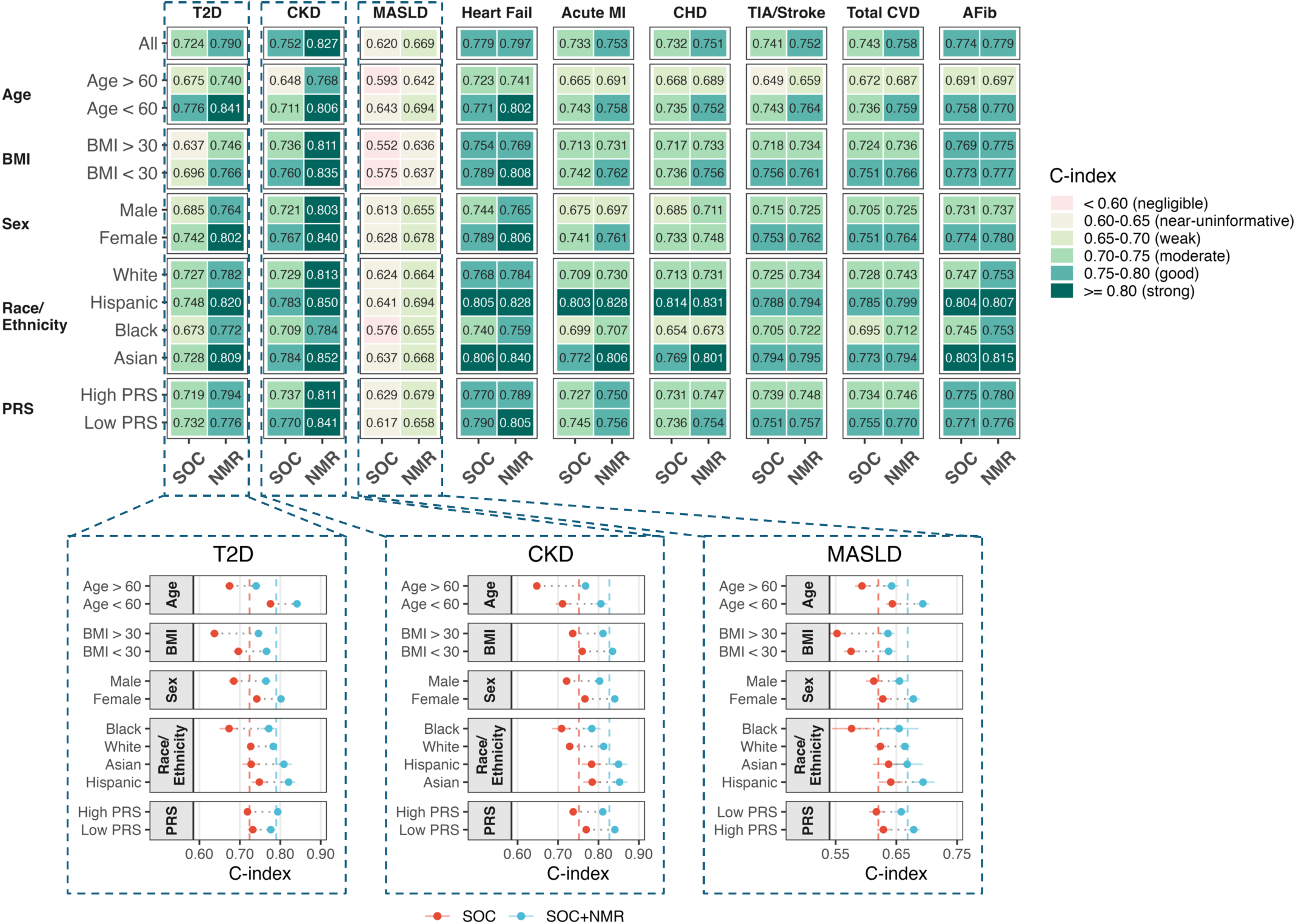
NMR metabolomics improves risk discrimination in subgroups where standard-of-care prediction is weakest. Top, heatmaps show C-indices for SOC and SOC+NMR models across all nine disease endpoints, evaluated overall and within strata defined by age, BMI, sex, race/ethnicity, and disease-specific polygenic risk. Within-stratum evaluation assesses discrimination among patients with similar characteristics while reducing the influence of between-group differences in baseline risk. Bottom, expanded views for T2D, CKD, and MASLD show subgroup-specific C-indices for SOC and SOC+NMR models. Points and whiskers indicate C-indices and 95% confidence intervals; dashed vertical lines indicate the corresponding overall-cohort C-indices for each model.

This pattern was clearest for T2D. SOC discrimination varied widely across patient groups, with C-indices ranging from approximately 0.64 to 0.78, and was lowest among participants with obesity (0.637), Black participants (0.673), men (0.685) and older participants (0.675), despite a cohort-wide SOC C-index of 0.724. Adding NMR improved discrimination in all 12 strata and produced substantially more uniform performance, with C-indices of approximately 0.74–0.84. Only two of 12 strata reached a C-index of 0.75 with SOC alone, compared with 11 of 12 with SOC+NMR; moreover, SOC+NMR met or exceeded the overall SOC benchmark in every stratum.

Similar compensation for weak subgroup-specific SOC performance was observed for CKD and MASLD. For CKD, SOC discrimination was lowest in older and Black participants, at approximately 0.65–0.71, whereas SOC+NMR raised discrimination across all patient groups to approximately 0.77–0.85, with the largest gains in the weakest-performing strata. For MASLD, NMR improved every subgroup comparison, including among participants with obesity (from 0.555 to 0.636) and Black participants (from 0.580 to 0.655), although absolute discrimination remained more modest overall.

These subgroup gains also translated into stronger enrichment of observed events among participants at highest predicted risk, particularly for T2D and CKD (Extended Data Fig. 3). For T2D, adding NMR increased 5-year incidence in the subgroup-specific top 5% by 1.5- to 2.1-fold—including from 25.8% to 53.7% among Black participants, and from 33.3% to 63.0% in participants with obesity—while calibration was generally preserved or improved across subgroups, with endpoint-specific exceptions, most notably for CKD.

### 2.4 Polygenic risk scores do not improve risk stratification beyond NMR metabolomics

Polygenic risk scores are increasingly being considered for integration into clinical risk assessment, based on their potential to identify inherited susceptibility before conventional risk factors become apparent^30–33^. Their practical value, however, depends on whether they improve prediction beyond the clinical information already available at the point of care. We therefore assessed whether endpoint-specific polygenic risk scores improved prediction across the nested clinical models and directly compared their added value with that of NMR metabolomics. We curated 62 published PRSs spanning the primary disease areas and closely related traits, including multiple candidate scores per endpoint (Supplementary Table 8). For each endpoint, we then selected the best-performing PRS, deliberately favoring PRS performance and thereby making the comparison conservative for detecting greater incremental predictive value of NMR (Supplementary Table 9).

The selected scores improved discrimination beyond age and sex alone, most clearly for T2D, for which the C-index increased from 0.560 to 0.626 (Extended Data Fig. 4, Supplementary Table 10). This added value diminished as more clinical information was incorporated. At the SOC level, across T2D, CKD and MASLD, adding PRS increased the C-index by only 0.007–0.02, compared with 0.05–0.08 for adding NMR to the same models. This pattern extended across endpoints, and PRS produced little meaningful further improvement once NMR metabolomics was included. Thus, although informative in relatively sparse baseline models, PRS did not materially improve clinically anchored risk stratification beyond contemporaneous clinical and metabolomic information.

### 2.5 NMR metabolomics improves prediction of disease progression and secondary outcomes

For patients already diagnosed with cardiometabolic diseases, the clinical question shifts from predicting disease onset to identifying those most likely to experience disease progression, including complications and other adverse post-diagnosis outcomes. Progression across the CKM continuum often extends beyond the initially affected organ: diabetes increases the risk of kidney disease and heart failure, while chronic kidney disease increases the risk of heart failure and coronary disease^4^. We therefore tested whether NMR metabolomics improved prediction of disease progression among participants with prevalent type 2 diabetes, chronic kidney disease or MASLD at enrollment, the three diseases with the largest number of secondary outcomes represented in the dataset.

We excluded participants with prior evidence of the secondary outcome being predicted. For these post-diagnosis analyses, BASE models included age, sex and disease duration, defined as the interval from first recorded diagnosis to serum sample collection; disease duration was therefore included in all subsequent nested models. CLIN models additionally included body mass index and systolic blood pressure, while disease-specific “SOC+” models further incorporated smoking status, outcome-relevant laboratory and treatment information, and measures of prior disease course, including HbA1c variability for diabetes, kidney-function trajectory for CKD and the NAFLD fibrosis score for MASLD (see Methods for details).

NMR improved prediction of disease progression across T2D, CKD, and MASLD (Fig. 4a,b; Extended Data Fig. 5, Supplementary Table 11 Supplementary Table 12). In participants with T2D, the largest improvements were observed for incident CKD (ΔC-index +0.08), broad renal disease (+0.06), and heart failure (+0.03). Among participants with CKD, NMR improved prediction of both heart failure and coronary heart disease (+0.03 each). In MASLD, the largest improvement was observed for portal hypertension (+0.20), although this estimate was based on relatively few events (Extended Data Fig. 5). Across all outcomes, CLIN+NMR generally matched or exceeded SOC+, with the exception of retinopathy. High-risk enrichment increased accordingly—for example, in the top-1% of predicted risk, 5-year incidence rose from 51.7% to 87.9% for CKD in diabetes and from 8.82% to 27.5% for portal hypertension in MASLD— while calibration was largely preserved or improved (Supplementary Fig. 9, Supplementary Fig. 10).

**Figure 4.**
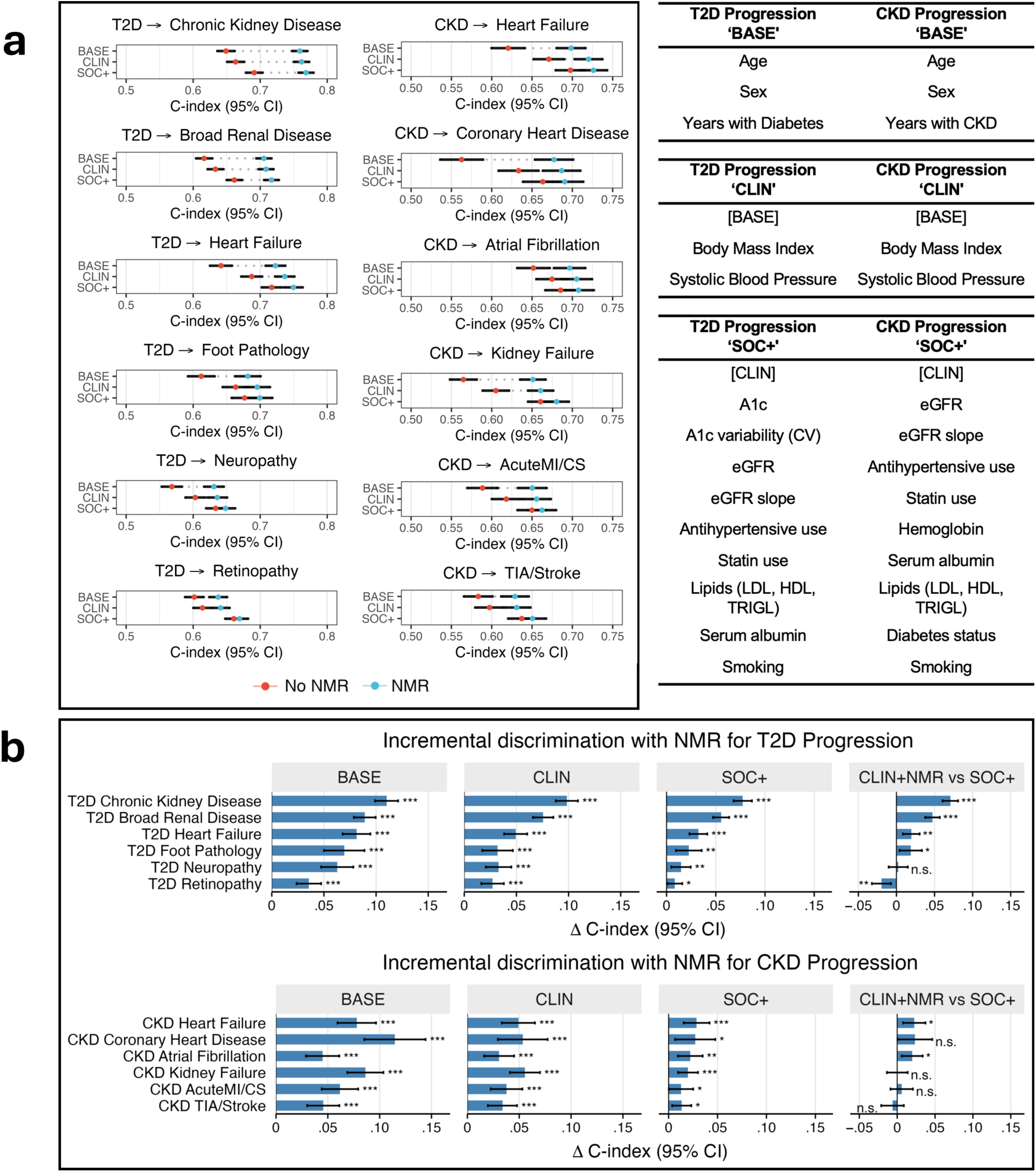
NMR metabolomics improves disease progression risk stratification after diabetes or chronic kidney disease diagnosis. Prediction of incident complications among participants with prevalent type 2 diabetes or chronic kidney disease at the time of NMR profiling. MASLD results are in Extended Data Fig. 5. **a,** Model discrimination for clinical risk models with and without the 250-biomarker NMR panel across diabetes- and CKD-related complications. **b,** Change in discrimination after adding NMR to each clinical model. Error bars indicate 95% confidence intervals. * p<0.05, **p<0.01, ***p<0.001, for FDR-corrected p-values.

### 2.6 NMR-enhanced models improve risk-based clinical prioritization

Improved discrimination and better enrichment of high-risk individuals are important, but they are not sufficient on their own to justify clinical use. For a risk model to be useful in practice, better ranking of patients should translate into better clinical decisions. In decision-curve analysis^34,35^, a “risk threshold” marks the predicted probability at which the expected benefit of further clinical action is judged to outweigh the consequences of unnecessary action. We used this approach to assess whether adding NMR-based risk estimates improved clinical prioritization across risk thresholds that might prompt closer follow-up, confirmatory testing, preventive counseling, or disease-specific surveillance. The net benefit then captures the balance between correctly identifying people who will go on to develop disease and unnecessarily flagging people who would remain healthy.

We illustrate this framework using incident type 2 diabetes, for which elevated predicted risk could prompt concrete preventive actions (Fig. 5a). At a representative 20% predicted 5-year risk threshold, a clinician might consider earlier follow-up, repeated HbA1c testing, prevention counseling, or review of cardiometabolic treatment. For context on the choice of this threshold, T2D prevention is recommended for people with prediabetes^36–38^, among whom reported 5-year progression risks can be well below 20%^4^; cardiovascular guidelines similarly consider preventive therapy at 10-year ASCVD risks below 20%^39,40^. At this threshold, the SOC model provided a net benefit equivalent to 6.60 correctly prioritized patients per 1,000 eligible patients, compared with 21.6 per 1,000 for the SOC+NMR model. Thus, using SOC+NMR yielded the equivalent of 15 additional true-positive prioritizations per 1,000 eligible patients for preventive diabetes care, while maintaining the same 20% decision threshold and therefore the same tolerance for unnecessary follow-up.

**Figure 5.**
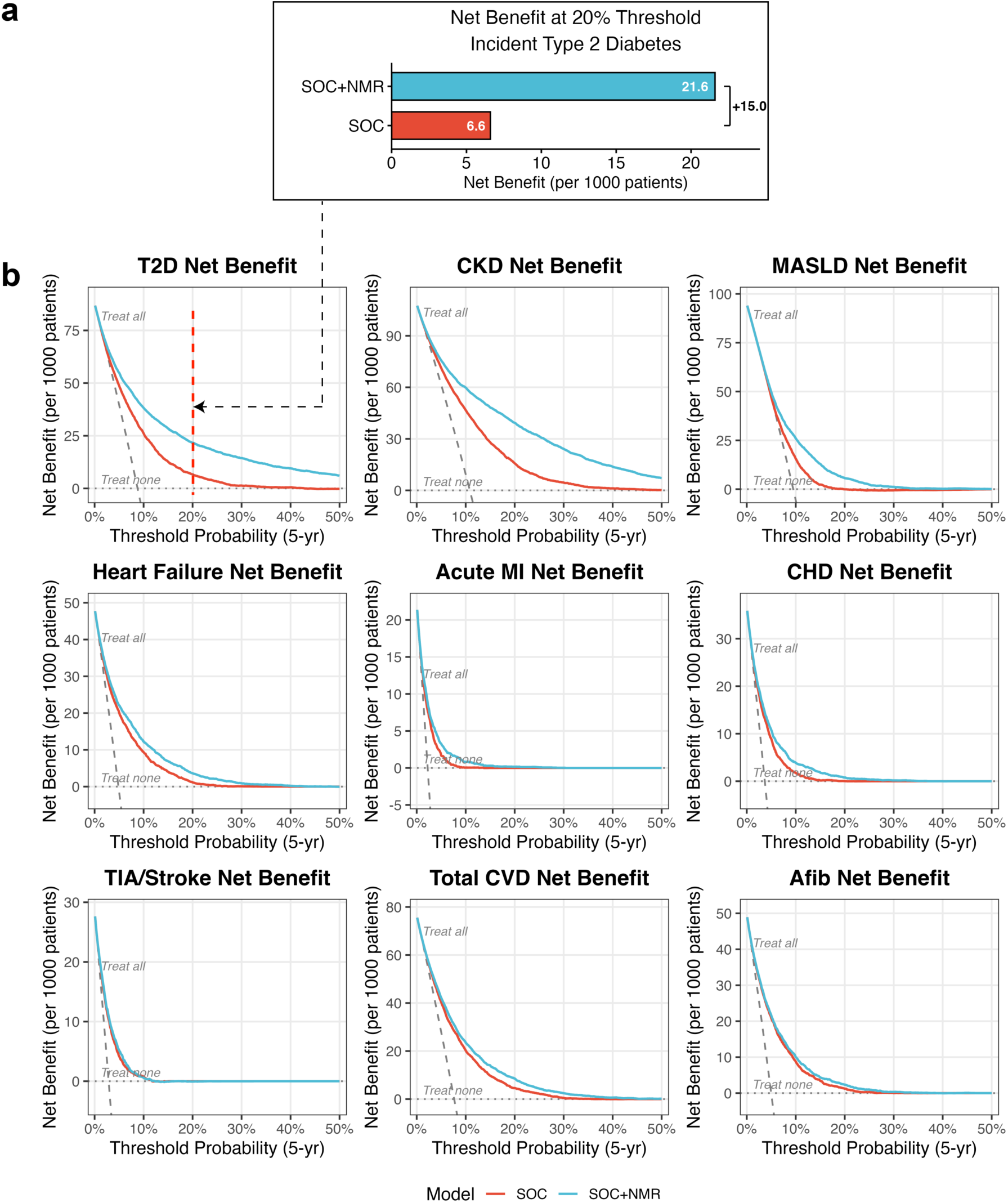
NMR-enhanced models improve clinical net benefit across risk thresholds. **a,** Net benefit for incident type 2 diabetes at a 20% predicted 5-year risk threshold, expressed as net true-positive-equivalent prioritizations per 1,000 eligible participants. Net benefit increased from 6.6 per 1,000 with SOC to 21.6 per 1,000 with SOC+NMR, corresponding to 15 additional correctly prioritized future cases per 1,000 at the same decision threshold. **b,** Decision-curve analysis across nine incident disease endpoints using out-of-fold 5-year risk predictions. SOC+NMR generally provided greater net benefit than SOC across the evaluated risk thresholds, with the largest gains for type 2 diabetes, chronic kidney disease, and MASLD.

We then extended this decision-curve evaluation across all nine incident disease endpoints (Fig. 5b; Supplementary Table 13). Across the evaluated ranges of predicted 5-year risk, SOC+NMR models generally provided higher net benefit than SOC models. The largest and most sustained gains were observed for T2D, CKD, and MASLD, consistent with the endpoints for which NMR produced the strongest improvements in discrimination and high-risk enrichment. Smaller and more threshold-dependent gains were observed for several cardiovascular outcomes. Taken together, the additional information captured by NMR metabolomics improved not only the ordering of patients by risk, but also the potential clinical utility of selecting patients for further assessment or preventive action.

### 2.7 Patient-level reports illustrate the clinical presentation of NMR-enhanced risk estimates

We next examined how the cohort-level gains in clinical prioritization could translate to a clinician-facing format for individual patients. We generated reports for three KPRB participants who were free of T2D, CKD or MASLD at enrollment and subsequently developed the corresponding disease (Fig. 6). The examples were selected using a common set of criteria: predicted 5-year risk was below the illustrative 20% threshold with the SOC model but above after addition of NMR, endpoint-relevant routine biomarker measurements were available both before and after enrollment, and the incident diagnosis occurred more than one year later. This yielded examples in which NMR changed risk prioritization well before the recorded diagnosis and in the context of longitudinal routine clinical information. The reports show how these risk estimates could be presented alongside the clinical trajectory in which they would be interpreted and are not intended to represent all participants with elevated NMR-enhanced risk.

1. In the report for the individual who developed T2D (Fig. 6, left panel), the patient was a 30-35 year-old woman with class III obesity and borderline prediabetes-range HbA1c who was normotensive and non-diabetic at baseline. The SOC estimate put her at a 17.6% risk of developing diabetes at the 5-year mark, compared to an estimated 39.0% risk based on the SOC+NMR model (∼2.2-fold higher). During follow-up, this participant developed hypertension and was diagnosed with type 2 diabetes approximately 3.3 years after the baseline blood draw, with metformin initiated around the time of diagnosis.
2. In the chronic kidney disease report (Fig. 6, middle panel), a 60-65 year-old woman with obesity and newly diagnosed hypertension had almost normal kidney function at baseline, with an eGFR near the reference range— a finding that would not, on its own, suggest markedly elevated CKD risk. The SOC model estimated 5-year CKD risk at 8.8%, whereas the SOC+NMR model estimated a substantially higher risk of 51.0% (∼5.8-fold higher). During follow-up, eGFR progressively declined, and CKD was diagnosed approximately 4.6 years after the baseline blood draw.
3. In the MASLD report (Fig. 6, right panel), the participant was a 55-60 year-old woman with obesity and only mildly out-of-range FIB-4 at baseline, a profile in which routine fibrosis-related markers alone would not have suggested markedly elevated near-term MASLD risk. The SOC model estimated a 5-year MASLD risk of 14.6%, whereas SOC+NMR estimated a substantially higher risk of 46.6% (∼3.2-fold higher). During follow-up, the participant’s FIB-4 rose steeply into the high-risk range, and MASLD was diagnosed approximately 2.2 years after the baseline blood draw. The participant later received furosemide and spironolactone, consistent with treatment for fluid overload, and died approximately six months after diagnosis.

**Figure 6.**
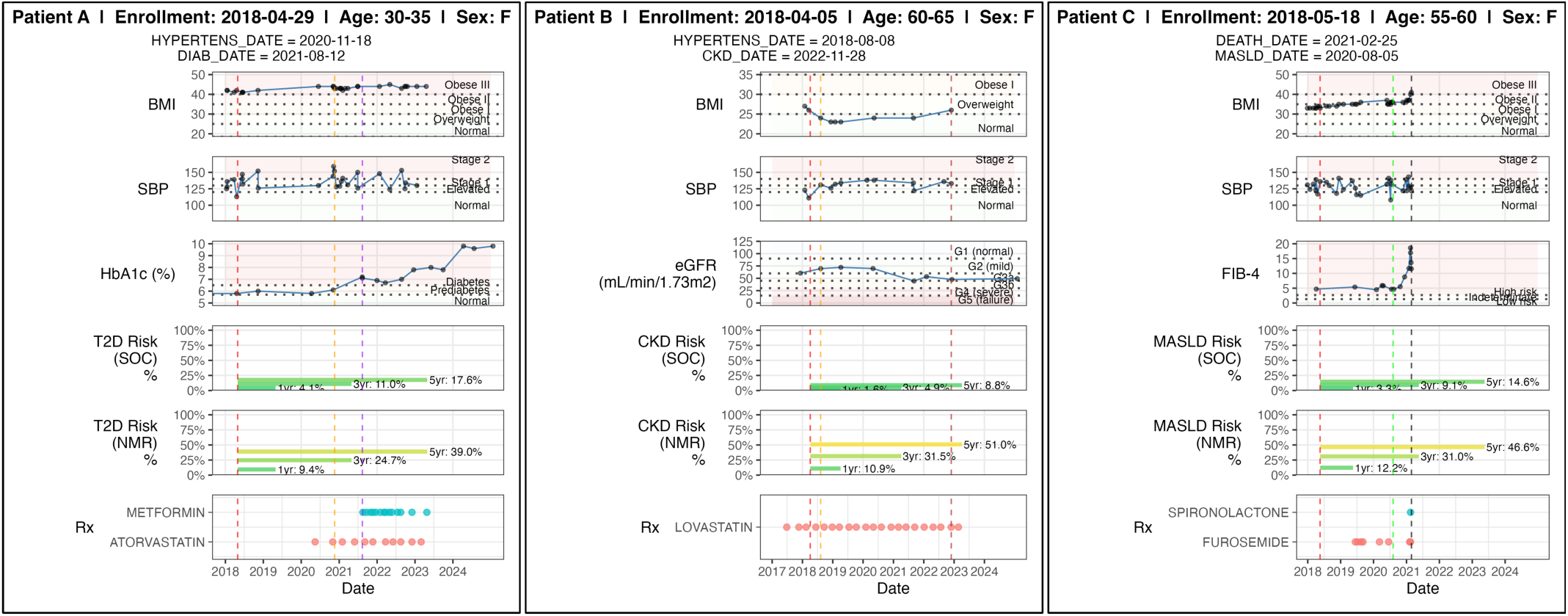
Examples of three patient-level NMR risk reports for incident type 2 diabetes (left panel), CKD (middle panel), and MASLD (right panel). Representative patient-level reports showing longitudinal EHR trajectories and predicted disease risk at the time of KPRB blood sample collection. Each panel shows body mass index, systolic blood pressure, endpoint-relevant clinical biomarkers, medication dispensing history where available, and 1-, 3- and 5-year predicted risk from SOC and SOC+NMR models. Vertical lines mark longitudinal EHR events: red, KPRB enrollment and blood draw; yellow, first hypertension diagnosis; purple, first type 2 diabetes diagnosis; brown, first CKD diagnosis; green, first MASLD diagnosis; and black, death. Dates shown were randomly shifted for de-identification, with relative timing between events preserved.

Together, these examples illustrate how NMR-enhanced risk estimates could be integrated with longitudinal clinical information in a clinician-facing report. They demonstrate the presentation and interpretation of model output, whereas the cohort-level analyses from the previous sections provide evidence for predictive performance and potential clinical utility.

## 3 Discussion

Population-based biobank studies have linked targeted NMR metabolomic profiles to future disease and demonstrated multi-disease risk prediction^14–16,41–43^. Whether this predictive value extends to routine healthcare, where measurements are irregular, incomplete, and shaped by healthcare use^23–25,44^, has remained uncertain. In 54,933 participants from a diverse, integrated US healthcare system, NMR improved risk stratification beyond routine clinical information. Importantly, predictive value extended beyond disease onset to post-diagnosis risk stratification, including disease-specific and cross-organ complications, spanning primary and secondary prevention.

The largest gains for incident disease were observed for T2D, CKD, and MASLD, with smaller improvements across most cardiovascular outcomes. This pattern may partly reflect overlap with the standard-of-care predictors: PREVENT already incorporates total and HDL cholesterol, which overlap directly with the NMR lipoprotein panel, whereas HbA1c, eGFR and FIB-4 are largely distinct from the NMR measurements, making the NMR profile more complementary for T2D, CKD and MASLD^8,13,29^. The stronger gains for metabolic and organ-function endpoints are consistent with prior biobank studies^14,15^. Among cardiovascular endpoints, atrial fibrillation showed essentially no additional discrimination beyond CHARGE-AF, while Total CVD necessarily averages across heterogeneous mechanisms. PRS added little beyond routine clinical predictors. Strikingly, a model combining NMR with only basic clinical measures outperformed the full disease-specific SOC models for eight of nine endpoints and performed similarly for atrial fibrillation. This suggests that a single NMR assay could potentially replace the different laboratory tests used for disease-specific risk assessment.

One potential concern is that the real-world setting itself could inflate the apparent benefit of NMR. Because routine clinical predictors are measured irregularly and their availability depends partly on patients’ healthcare engagement, SOC models may be disadvantaged relative to a standardized biobank setting^44^. However, similar NMR gains in complete-case and healthcare-engagement sensitivity analyses argue against this explanation. Excluding diagnoses within six months of sampling also reduced the possibility that NMR was capturing occult disease already present at baseline rather than future disease risk. Finally, when models developed in UK Biobank were evaluated in KPRB, overall discrimination was lower, but an incremental NMR contribution was retained for several endpoints, most clearly T2D, CKD and MASLD. Together, these analyses argue that the NMR contribution observed in KPRB is not merely an artifact of the routine-care setting, while also showing that biobank-derived risk models may not generalize unchanged across populations and settings.

After diagnosis, the clinical question shifts to identifying patients at greatest risk of adverse outcomes. Among patients with established T2D, CKD, or MASLD, NMR added prognostic information beyond disease duration, current clinical measurements, treatment information, and longitudinal markers of disease control and progression. This extended to disease-related complications—renal complications in T2D, kidney failure in CKD and portal hypertension in MASLD—as well as cross-organ outcomes, including heart failure in T2D and heart failure and coronary heart disease in CKD. Notably, several of the strongest gains in discrimination were observed in these post-diagnosis analyses, including for renal outcomes in T2D and liver disease progression in MASLD. The ability to stratify risk for such cross-organ outcomes is particularly relevant to CKM progression, which frequently extends beyond the initially affected organ^1^. Prior metabolomic studies have examined prognosis within individual diseases, including cardiovascular events in type 2 diabetes^45,46^, diabetic kidney disease^47,48^, cardiovascular events in chronic kidney disease^49^, and liver-related outcomes in MASLD^50^. Our findings extend these observations across multiple established diseases and complications, including both within-organ and cross-organ outcomes, supporting a broader role for NMR in ongoing disease management and secondary prevention.

For NMR-based risk stratification to be useful across clinical populations, its added value should not be confined to particular patient groups. Cardiometabolic risk models can perform unevenly across demographic and anthropometric groups, while overall discrimination can obscure weaker performance within them^9,51^. In KPRB, NMR improved discrimination across nearly all age, BMI, sex, race-and-ethnicity strata, and polygenic-risk strata, with the largest gains often occurring where SOC performed least well. This within-group performance is important for clinical translation because it suggests that the observed NMR benefit is not merely a consequence of population heterogeneity and may be most useful where conventional risk prediction is weakest.

Ultimately, more consistent risk stratification only becomes clinically useful when it translates into better patient prioritization in practice. In KPRB, NMR-enhanced risk estimates were generally well calibrated, concentrated more future cases among the patients prioritized as highest risk, and provided greater net benefit across clinically plausible thresholds, allowing the clinical benefit to be quantified at different tolerances for unnecessary follow-up. For T2D, at a 20% predicted 5-year risk threshold, NMR corresponded to approximately 15 additional future cases correctly prioritized per 1,000 eligible patients after accounting for unnecessary follow-up. This could help prioritize higher-risk patients for further assessment and, where prediabetes is confirmed, established preventive interventions^36–38^. Patient-level reports further illustrate how calibrated NMR-enhanced risk estimates could be interpreted alongside longitudinal clinical information. Together, these findings suggest that NMR may improve not only risk estimation but also clinical prioritization.

This study has several limitations. Disease onset and complications were defined from longitudinal EHR data and therefore reflect recorded clinical recognition rather than biological onset, with diagnosis timing partly dependent on healthcare contact and evaluation. The KPRB metabolomics cohort was enriched for selected disease areas, longer follow-up, and racial and ethnic diversity, rather than randomly sampled from the broader KPRB population, so absolute event rates and risk estimates may not represent the broader KPRB population. Although UK Biobank–trained models retained the added contribution of NMR when evaluated in KPRB, their lower overall performance indicates that fitted models may require local validation, recalibration, or refitting. Some post-diagnosis analyses, particularly in MASLD, included fewer events and yielded less precise estimates. Finally, decision-curve benefit supports potential clinical utility but does not establish that NMR-informed risk assessment changes care or improves patient outcomes, which will require prospective evaluation.

Together, these findings support targeted NMR metabolomics as a common risk-stratification approach across chronic diseases, both before and after diagnosis, in an integrated healthcare system. The KPRB metabolomics resource generated here provides a platform for further biological discovery and translational research. Prospective studies are now needed to determine how locally calibrated NMR-based risk estimates can be integrated into primary and secondary prevention and whether their use improves clinical decision-making and patient outcomes.

## 4 Methods

### 4.1 Kaiser Permanente Research Bank cohort

The Kaiser Permanente Research Bank (KPRB) is a longitudinal research biobank embedded within the Kaiser Permanente integrated healthcare system, with participant biospecimens and survey data linked to electronic health record (EHR) data^28^. The broader KPRB comprises approximately 440,000 adult participants^28^. For the present study, one banked serum sample from each of 54,933 KPRB participants was selected for nuclear magnetic resonance (NMR) metabolomic profiling. Samples were collected between April 11, 2015 and September 10, 2021, and the date of collection of the serum sample used for NMR profiling was defined as the study index date. Fasting status was not uniformly available for KPRB serum samples. The metabolomics subcohort was selected to support racial and ethnic diversity, longer available longitudinal EHR follow-up and enrichment for cardiometabolic, kidney and pulmonary disease. The sampling strategy and participant flow are summarized in Extended Data Fig. 1.

All recruitment and analyses were reviewed and approved by the Kaiser Permanente Institutional Review Board (Protocols 1283755-470 and 2052704-28). All KPRB participants provided written informed consent at enrollment, including consent for use of their medical records, survey, and biospecimen data in research.

### 4.2 NMR metabolomic profiling and preprocessing

Serum samples were profiled using the Nightingale Health ^1^H nuclear magnetic resonance (NMR) metabolomics platform^14,16,29^. The platform quantifies circulating biomarkers spanning size-resolved lipoprotein particle concentrations and lipid composition, including triglycerides, cholesterol and phospholipids; fatty acids; amino acids; glycolysis-related metabolites; ketone bodies; other small molecules; and inflammation-related protein markers. Glycerol was excluded because of high missingness, leaving 250 NMR biomarkers, including derived measures and ratios, for downstream analyses. All retained biomarkers were log2-transformed before analysis. Biomarker definitions, Nightingale nomenclature, units, limits of detection, distributions and missingness are provided in Supplementary Table 4.

Needle-to-freezer time, defined as the interval in hours between blood collection and sample freezing, introduced systematic pre-analytic variation in the KPRB NMR measurements and was therefore accounted for before downstream analysis. For each NMR biomarker, the association with needle-to-freezer time was quantified using Pearson correlation. Associations were observed for nearly all biomarkers, although effect sizes were generally small, with most absolute correlation coefficients below 0.10 and only a small number approaching 0.20. Each biomarker was residualized against needle-to-freezer time, and the resulting residuals were recentered using the median abundance among samples frozen within 0–32 h to preserve the original biomarker scale. The corrected biomarker values were used in all subsequent KPRB analyses. Detailed needle-to-freezer associations and the effect of the correction are shown in Supplementary Fig. 1.

### 4.3 Disease outcome definitions and ascertainment

Nine individual disease endpoints were prespecified before model fitting and evaluation: type 2 diabetes (T2D), chronic kidney disease (CKD), metabolic dysfunction-associated steatotic liver disease (MASLD), heart failure including cardiomyopathy, acute myocardial infarction (MI), coronary heart disease (CHD), (TIA)/stroke, Total CVD (see below), atrial fibrillation (Afib). Chronic obstructive pulmonary disease (COPD) was used as an additional non-CKM outcome. For the CKM-focused analyses, Total Cardiovascular Disease (Total CVD) was additionally evaluated as a composite of CHD, acute MI, TIA/stroke and heart failure; COPD was evaluated separately as a pulmonary endpoint. MASLD ascertainment included related historical non-alcoholic fatty liver disease terminology^52^. Complete ICD-10 code-based definitions are provided in Supplementary Table 14. CHD was furthermore defined by procedural CPT codes that indicate heart disease, in addition to ICD-10 coding.

Secondary outcomes were defined for participants with T2D, CKD or MASLD and included renal and cardiovascular complications, retinopathy, neuropathy and diabetic foot disease for T2D; kidney failure/end-stage renal disease and cardiovascular outcomes for CKD; and liver disease progression, portal hypertension and deep vein thrombosis for MASLD. Complete definitions, ICD-10 codes, and event counts are provided in Supplementary Table 15.

### 4.4 Clinical data and baseline feature construction

Clinical information was obtained from longitudinal electronic health record (EHR) data linked to KPRB^28^. Variables used in the analyses included age at serum sample collection, self-reported sex, self-reported race and ethnicity, height, weight, body mass index (BMI), systolic and diastolic blood pressure, HbA1c, serum creatinine, aspartate aminotransferase (AST), alanine aminotransferase (ALT), platelet count, total cholesterol, HDL cholesterol, LDL cholesterol, triglycerides, albumin, current smoking status, diabetes status, antihypertensive medication use, lipid-lowering medication use, and history of myocardial infarction and heart failure. Medication use was determined from dispensing records, and prior disease status was determined from the longitudinal EHR. A detailed list of all recorded medications is shown in Supplementary Table 16. The cardiovascular and atrial-fibrillation variables reflect the component predictors used in the PREVENT-and CHARGE-AF–based predictor sets^8,53^.

Derived clinical measures included estimated glomerular filtration rate (eGFR^54^) and FIB-4. FIB-4 was calculated from age, AST, ALT, and platelet count^55,56^. These raw and derived clinical variables were used either directly as predictors or as components of the endpoint-specific clinical predictor sets described below. Definitions, units, derivations, baseline distributions, and missingness of the clinical variables, together with medication summaries, are provided in Supplementary Table 3.

Because clinical measurements were obtained during routine care rather than at a standardized study visit, their timing and availability varied across participants. BMI and systolic blood pressure, which were required covariates in all CLIN models, were routinely measured and had little overall missingness. For these variables, values at the index date were estimated by linear interpolation between the nearest available measurements before and after the index date. We used this approach because BMI and blood pressure would generally be expected to be available at the point of clinical application; thus, the absence of a measurement exactly at the index date primarily reflects the timing of routine clinical assessments rather than true unavailability of the variable.

For other routinely measured clinical variables, only information recorded on or before serum sample collection was considered, and the most recent measurement within the preceding two years was identified. In the primary KPRB analyses, each variable was represented by the most recent observed value, the time elapsed between that measurement and serum sample collection, and an indicator denoting whether the variable was unavailable within the two-year lookback window^23^.

### 4.5 Prediction models and internal validation

Separate time-to-event models were fitted for each disease endpoint. For incident-disease analyses, participants with evidence of the corresponding disease on or before serum sample collection were excluded. Follow-up continued from serum sample collection until the first qualifying endpoint event, death, Kaiser Permanente membership disenrollment or the end of available EHR follow-up, whichever occurred first; death before the endpoint of interest was treated as censoring. For secondary-outcome analyses, participants were required to have the underlying disease at serum sample collection and were excluded if the complication being predicted had already occurred.

For incident-disease prediction, three nested clinical predictor sets were defined. BASE included age at serum sample collection and sex; CLIN additionally included body mass index and systolic blood pressure; and standard-of-care (SOC) models further included endpoint-specific clinical risk information: HbA1c for type 2 diabetes, estimated glomerular filtration rate (eGFR) for chronic kidney disease, FIB-4 for metabolic dysfunction-associated steatotic liver disease, PREVENT-derived predictor sets for coronary heart disease, heart failure/cardiomyopathy, acute myocardial infarction and transient ischemic attack/stroke^8^, a CHARGE-AF–derived predictor set for atrial fibrillation^53^, and age, LDL cholesterol, and smoking status for chronic obstructive pulmonary disease. For PREVENT- and CHARGE-AF–based models, component predictors were entered individually and coefficients were re-estimated during model training. NMR-enhanced models additionally included all 250 retained NMR biomarkers. For secondary-outcome analyses, disease-specific SOC+ models incorporated disease duration, defined as the interval from first recorded diagnosis to serum sample collection, current clinical measurements, smoking status, endpoint-specific laboratory and treatment information, and disease-history summaries derived from information recorded on or before serum sample collection, including HbA1c variability for type 2 diabetes, eGFR trajectory for chronic kidney disease and the NAFLD fibrosis score for metabolic dysfunction-associated steatotic liver disease^57^. Complete SOC+ definitions and lookback windows are provided in Supplementary Table 17.

All models were fitted using ridge-penalized Cox regression in R v4.2.2 with `glmnet` v4.1.8 (`family = “cox”`, `alpha = 0`) ^58^. Ridge regularization was used because the predictor sets contained correlated clinical and metabolomic variables, particularly within the NMR panel Supplementary Fig. 2, allowing all 250 biomarkers to be modeled jointly while shrinking coefficient estimates to reduce variance and limit overfitting. Age and sex were assigned a penalty factor of 0.1 and all other predictors a penalty factor of 1.

Internal (within-KPRB) validation used five-fold cross-validation. For each endpoint, models were trained on four folds and applied to the remaining held-out fold until every participant had an out-of-sample prediction. All preprocessing and tuning were performed within the training partition. Continuous predictors were standardized using training-fold means and standard deviations and applied unchanged to the held-out fold; `glmnet` internal standardization was disabled. Categorical predictors were represented using indicator variables. Within each training partition, λ was selected by the minimum cross-validated error rule using concordance (`type.measure = “C”`) across 100 candidate values. Predictions from the five held-out folds were pooled for subsequent model evaluation. Thus, all reported internal-validation performance was based on predictions for participants who were not used for model fitting, preprocessing, or hyperparameter selection.

### 4.6 Polygenic risk score generation and analysis

Published polygenic scoring models relevant to the study endpoints were identified from the PGS Catalog^59^. A total of 62 candidate scores spanning the primary disease areas and closely related traits were manually curated. The complete set of candidate scores, including PGS Catalog identifiers, associated phenotypes, source publications and assignment to study endpoints, is provided in Supplementary Table 8.

Participant-level polygenic risk scores (PRS) were generated by KPRB using the PGS Catalog Calculator workflow^60^. For each candidate PGS Catalog identifier, the published scoring definition was retrieved from the PGS Catalog, genome-build compatibility was verified, and scoring variants were matched and harmonized to KPRB PMDA Version 2 TOPMed-imputed genotype data. Genotypes were represented as chromosome-level PLINK2^61^ v2.00a5.10 allele-dosage datasets on GRCh38, and dosage-weighted scores were calculated separately by chromosome and then aggregated to generate one participant-level score for each candidate PGS. The workflow was implemented using pgsc_calc^60^ v2.2.0, executed with Nextflow^62^ v24.10.9 in a containerized environment, and generated variant-matching summaries, quality-control reports and software-execution metadata.

For each disease endpoint, the best-performing PRS was selected from the candidate scores assigned to that endpoint. This deliberately favorable selection strategy was used to maximize PRS performance and thereby provide a conservative comparison of its incremental predictive value with that of NMR metabolomics. The selected PRS was represented as a continuous predictor and evaluated using the modeling and cross-validation framework described in Section 4.5. Comparisons included BASE versus BASE+PRS, SOC versus SOC+PRS, and SOC, SOC+PRS, SOC+NMR and SOC+PRS+NMR.

### 4.7 Model evaluation and statistical analyses

Model performance was evaluated using pooled held-out predictions from five-fold cross-validation. Discrimination was quantified using Harrell’s concordance index (C-index)^63^, and incremental performance was summarized as the difference in C-index (ΔC-index) between paired models. Primary comparisons were BASE+NMR versus BASE, CLIN+NMR versus CLIN, SOC+NMR versus SOC, and CLIN+NMR versus SOC. Paired C-index comparisons were performed using compareC v1.3.3 ^64^, with 95% confidence intervals calculated as the estimated difference ±1.96 standard errors from the asymptotic variance returned by compareC. Because the comparison of two concordance indices requires paired observations, all models within a comparison set were restricted to the intersection of participants with out-of-fold predictions, so that each paired test used an identical patient set and identical follow-up times. P values were adjusted across the nine disease endpoints within each model comparison using the Benjamini– Hochberg procedure, with FDR-adjusted P<0.05 considered statistically significant. Corresponding secondary-outcome and PRS comparisons were adjusted within their respective analysis sets.

Absolute 1-, 3- and 5-year risks were estimated from the cross-validated Cox models. Calibration was assessed by comparing predicted risk with Kaplan–Meier observed risk across deciles of predicted risk. High-risk enrichment was evaluated using observed 5-year incidence across the predicted-risk distribution and among participants in the top 1%, 5% and 10% of predicted risk. Subgroup performance was evaluated using held-out predictions within strata defined by age, body mass index, sex, self-reported race and ethnicity, and endpoint-specific polygenic risk; subgroup definitions and sample sizes are provided in Supplementary Table 7. Clinical utility was assessed using decision-curve analysis^34^, comparing SOC and SOC+NMR models across 5-year risk thresholds and expressing net benefit as net true-positive-equivalent prioritizations per 1,000 participants.

Three sensitivity analyses were performed for the incident-disease models. First, analyses were repeated among participants with complete clinical measurements within the two-year lookback window, with measurement-recency and missingness terms omitted. Second, events occurring within 6 and 12 months after serum sample collection were excluded to reduce the influence of disease that may already have been clinically unrecognized at baseline. Third, participants with no recorded EHR encounters during the two years preceding serum sample collection were excluded to assess the influence of limited recent healthcare-system engagement.

Patient-level reports for type 2 diabetes, chronic kidney disease and metabolic dysfunction-associated steatotic liver disease were generated as illustrative examples of how longitudinal clinical information and SOC versus SOC+NMR 1-, 3- and 5-year risk estimates could be presented together. These participants were selected to illustrate the report format and were not intended to represent the broader cohort. Dates shown in the reports were randomly shifted for de-identification, with relative timing between events preserved.

### 4.8 UK Biobank cohort, metabolomics and clinical endpoints

UK Biobank is a population-based prospective cohort of approximately 500,000 participants recruited across the United Kingdom between 2006 and 2010 at 40–69 years of age^65,66^. For the present study, analyses were restricted to participants with available NMR metabolomic measurements as of Aug 15, 2023 and the clinical and outcome data required for the corresponding endpoint-specific analyses (n = 248,006).

Demographic characteristics, anthropometric measurements, blood pressure and clinical chemistry measurements were collected using standardized procedures at the baseline assessment, with subsequent health outcomes ascertained through linkage to longitudinal health records^65,66^. NMR metabolomic profiling was performed in baseline EDTA plasma using the Nightingale Health platform^15^.

The same nine incident disease endpoints evaluated in KPRB were considered in UK Biobank: type 2 diabetes, chronic kidney disease, metabolic dysfunction-associated steatotic liver disease, coronary heart disease, heart failure including cardiomyopathy, acute myocardial infarction, transient ischemic attack/stroke, atrial fibrillation, and an additional supplemental chronic obstructive pulmonary disease outcome. Endpoint definitions were harmonized with those used in KPRB where applicable. For each endpoint, participants with evidence of the corresponding disease at or before baseline were excluded, and follow-up continued until the first qualifying endpoint event, death or the end of available linked follow-up. Complete endpoint definitions and cohort-specific follow-up characteristics are provided in Supplementary Table 2 and Supplementary Table 14.

UK Biobank operates under ethical approval from the North West–Haydock Research Ethics Committee (REC reference 21/NW/0157), and all participants provided written informed consent for research use of their data and biological samples^65^. The present analyses were conducted using the UK Biobank Resource under Application #588633.

### 4.9 External validation of UK Biobank–derived models in KPRB

To assess cross-cohort generalizability, endpoint-specific models were developed in UK Biobank and evaluated in eligible KPRB participants. The same endpoint definitions, eligibility criteria and BASE, CLIN, SOC and NMR-enhanced predictor sets described for the KPRB analyses were used. Because UK Biobank is a population-based cohort in which clinical measurements were collected systematically at the baseline assessment, the clinical predictors required for these analyses were largely complete. The KPRB-specific measurement-recency and missingness features were therefore not used in the UK Biobank models. Models were fitted using the same ridge-penalized Cox framework and tuning procedure described in the section above (*Prediction models and internal validation*).

NMR biomarkers were matched between cohorts by feature definition. After exclusion of glycerol, the 250 retained biomarkers were log2-transformed in both cohorts. UK Biobank measurements were obtained from plasma, whereas KPRB measurements were obtained from serum; serum and plasma are known to differ systematically in metabolite and lipoprotein concentrations ^67,68^. Consistent with these known matrix effects, systematic differences in NMR biomarker distributions between cohorts were observed in Supplementary Table 18 and Supplementary Fig. 11. Cross-cohort differences in NMR biomarker distributions were quantified using Cohen’s d. Each NMR biomarker was standardized separately within UK Biobank and KPRB to a mean of 0 and standard deviation of 1 before model development and evaluation. Model performance in KPRB was quantified using Harrell’s C-index^63^, and the incremental predictive value of NMR was assessed by comparing each NMR-enhanced model with its corresponding clinical model. This cross-cohort evaluation provided an additional test of whether the incremental predictive signal from NMR generalized beyond the cohort in which model coefficients were estimated.

## 5 Data Availability

Data from the Kaiser Permanente Research Bank (KPRB), including the NMR metabolomics and linked electronic health record data used in this study, are available to qualified researchers through the KPRB application process, subject to review and approval by KPRB and applicable institutional and data-use requirements. https://researchbank.kaiserpermanente.org/for-researchers/apply-for-access/. UK Biobank data are available to approved researchers through the UK Biobank access process. This study used UK Biobank data under Application #588633.

## 6 Code Availability

All code used to generate the figures and supplementary materials in this study is provided as R-based Jupyter notebooks on GitHub at https://github.com/krumsieklab/kprb-nmr-risk-strat.

## 9 Author Contributions

J.C.-B., S.R., O.E., K.S., D.A., and J.K. designed the study. G.R.V., S.R. and D.A. contributed to the KPRB cohort and data resources, and J.C.-B. and G.R.V. curated the clinical data. J.C.-B. and J.K. performed the computational and statistical analyses. J.C.-B., C.I., J.R., D.A. and J.K. interpreted the results. K.S., D.A. and J.K. supervised the work. J.C.-B., J.R., K.S., and J.K. wrote the manuscript. All authors reviewed and revised the manuscript.

---

**Extended Data Fig. 1:**
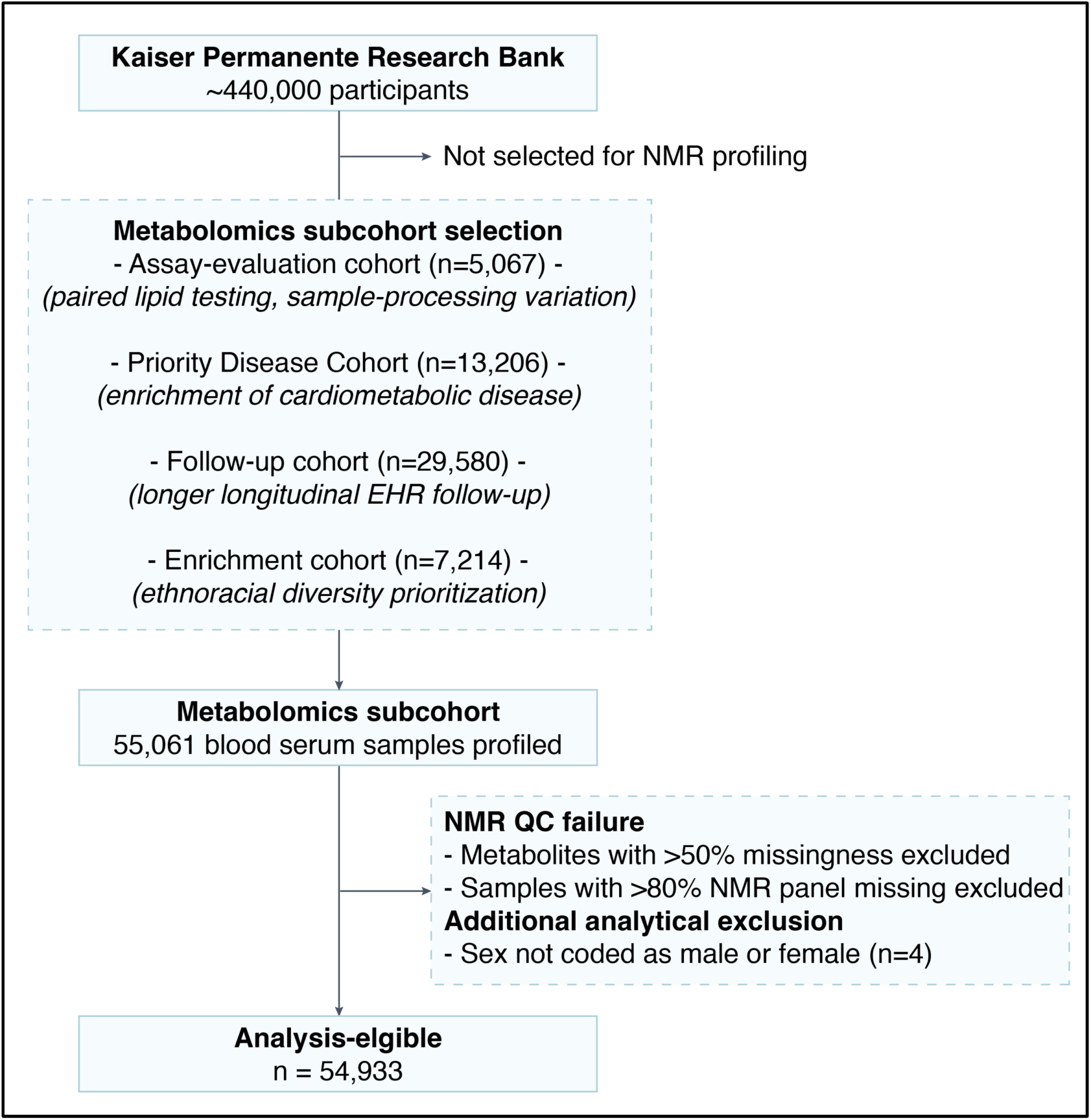
Selection of the KPRB metabolomics subcohort. Participant selection from the broader Kaiser Permanente Research Bank into four prioritized sampling groups: an assay-evaluation cohort (n=5,067), enriched for paired lipid testing within 60 days of enrollment and technical variation in sample processing; a priority disease cohort (n=13,206), enriched for cardiometabolic disease; a follow-up cohort (n=29,580), prioritized for longer longitudinal EHR follow-up; and an enrichment cohort (n=7,214), prioritized for ethnoracial diversity. Sampling was designed to represent approximately equal proportions of European and non-European ancestry while capturing variation in disease burden, follow-up duration and pre-analytic sample characteristics.

**Extended Data Fig. 2:**
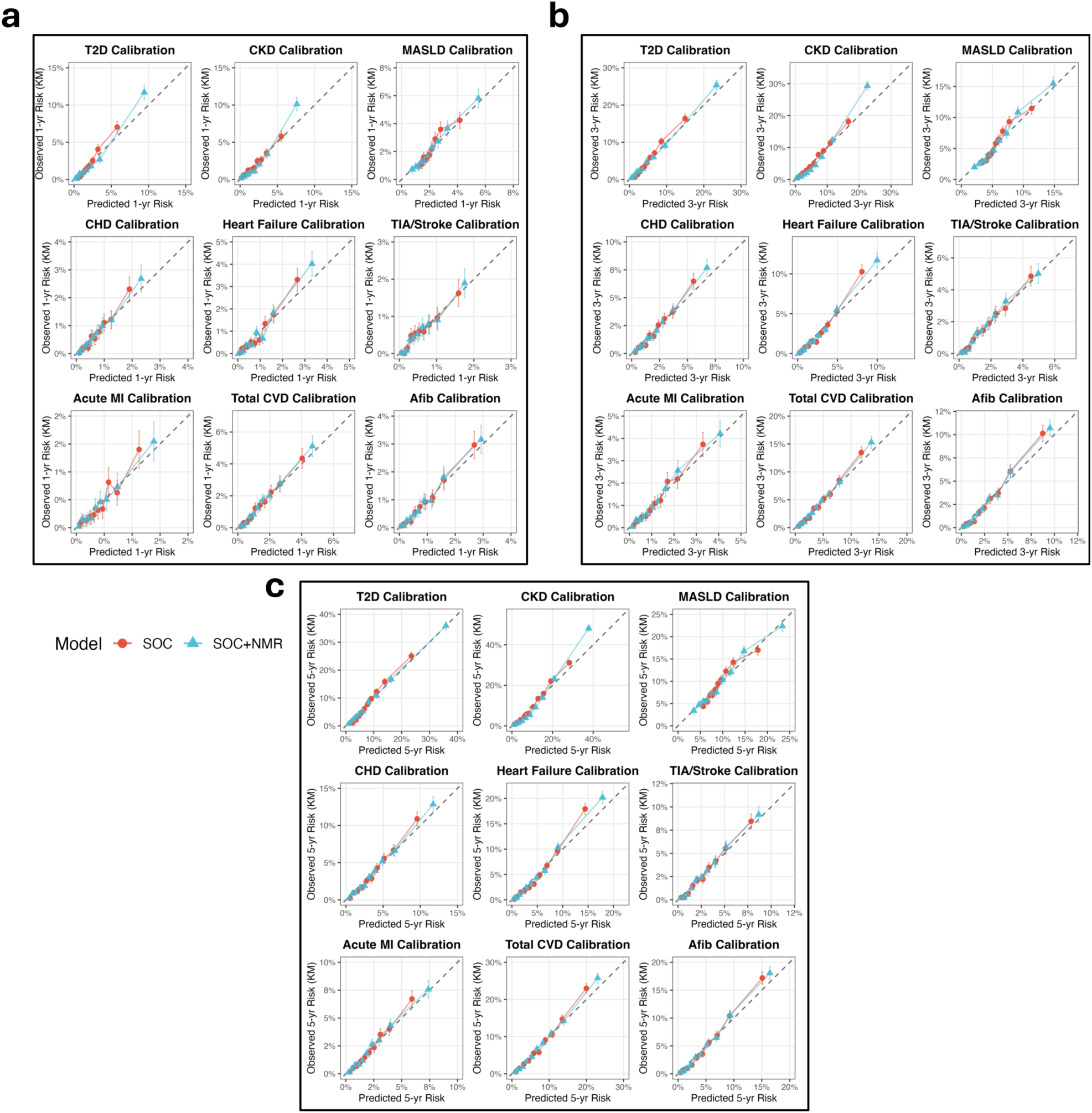
Calibration of incident disease risk models across prediction horizons. Observed and predicted risks across risk score deciles at **a,** 1, **b,** 3, and **c,** 5 years for SOC and SOC+NMR models across nine CKM endpoints. Across all time thresholds, model calibration was generally preserved or improved with SOC+NMR while expanding the dynamic range of predicted risk, although risk was slightly underestimated in the highest-risk decile for CKD.

**Extended Data Fig. 3:**
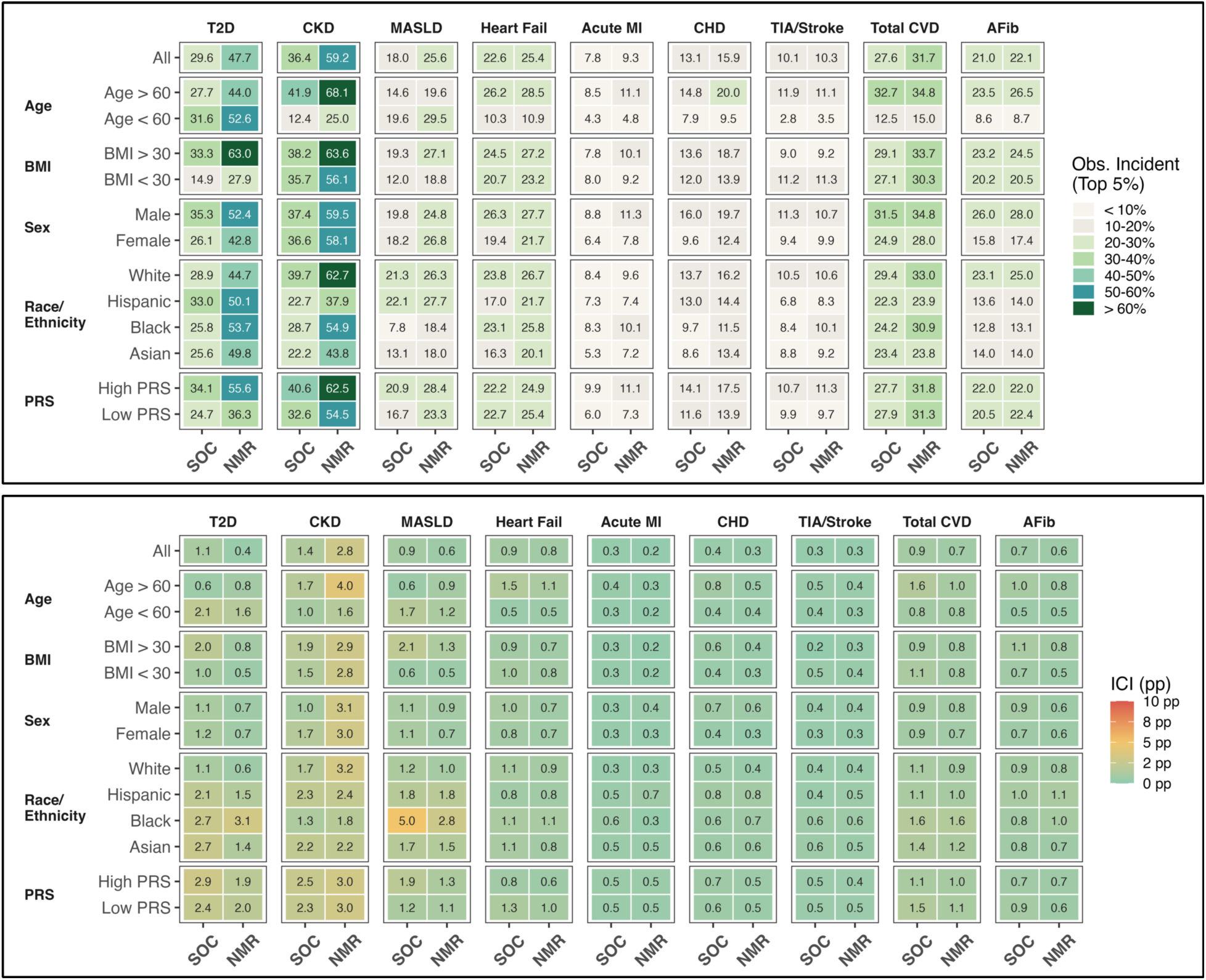
Subgroup-specific high-risk enrichment and calibration. High-risk enrichment and calibration across participant subgroups. Top, observed 5-year incidence among participants in the top 5% of predicted risk within each age, BMI, sex, race/ethnicity and PRS stratum for SOC and SOC+NMR models. Bottom, 5-year integrated calibration index (ICI), calculated as the n-weighted mean absolute difference between predicted and Kaplan–Meier–observed risk across equal-sized risk deciles. Lower ICI indicates better calibration, with 0 representing perfect agreement. The “All” row represents the full estimates obtained from the full cohort analysis. Across most subgroups, SOC+NMR increased 5-year incidence among the top 5% of predicted risk, with the largest enrichment gains for T2D, CKD and MASLD; gains were smaller for cardiovascular endpoints and minimal for TIA/stroke. Calibration was generally preserved or improved with NMR, although CKD showed consistently higher ICI across several strata.

**Extended Data Fig. 4:**
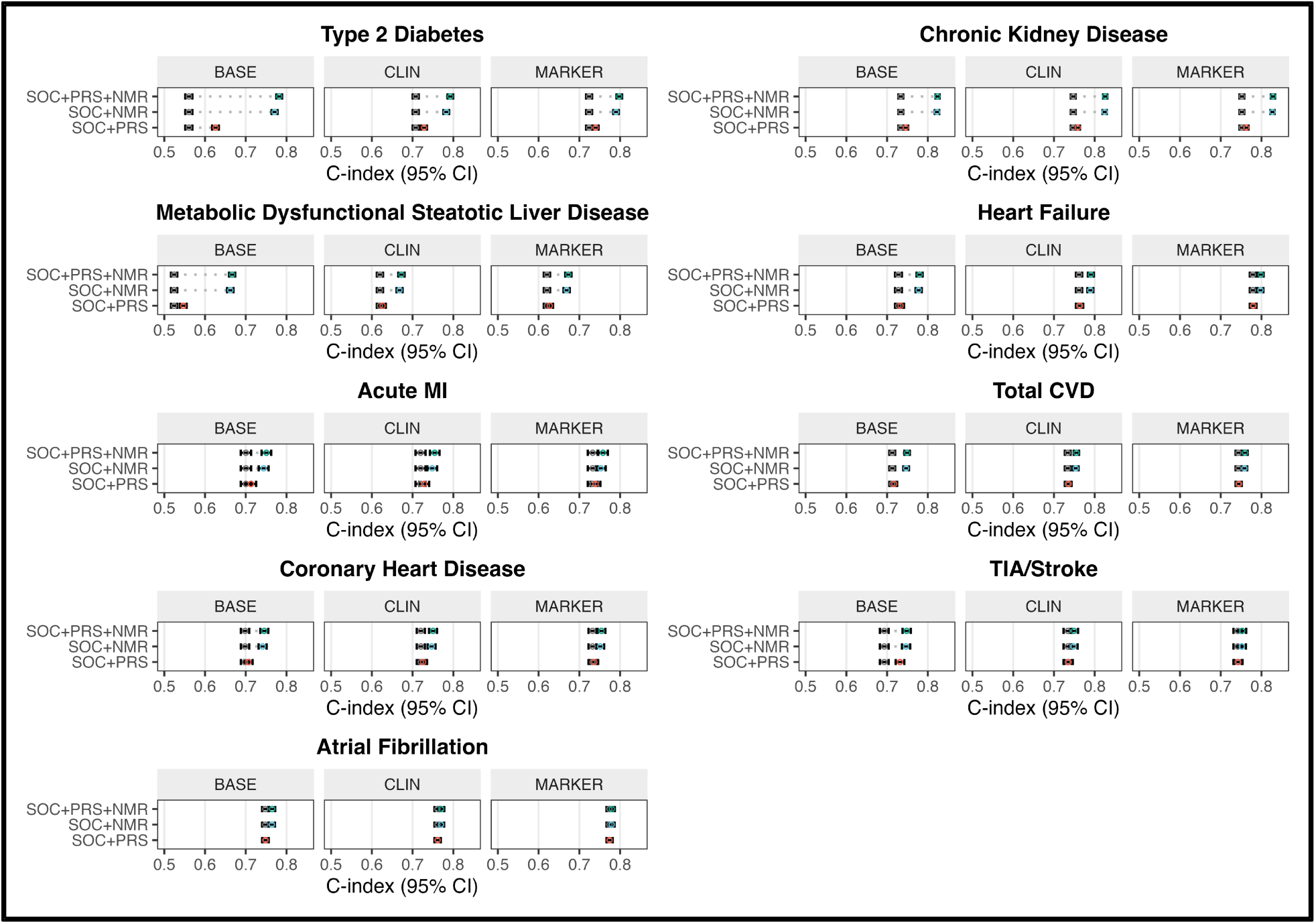
Incremental predictive value of polygenic risk scores and NMR metabolomics. C-index performance for models incorporating clinical predictors, endpoint-specific PRS and NMR metabolomics. For each endpoint, the best-performing PRS was selected from 62 curated published scores and evaluated across BASE, CLIN and MARKER predictor tiers. Points and error bars indicate C-index estimates and 95% confidence intervals for models with PRS, NMR, or both. PRS improved discrimination most in sparse baseline models, but its incremental contribution diminished with additional clinical information and was small once NMR was included.

**Extended Data Fig. 5:**
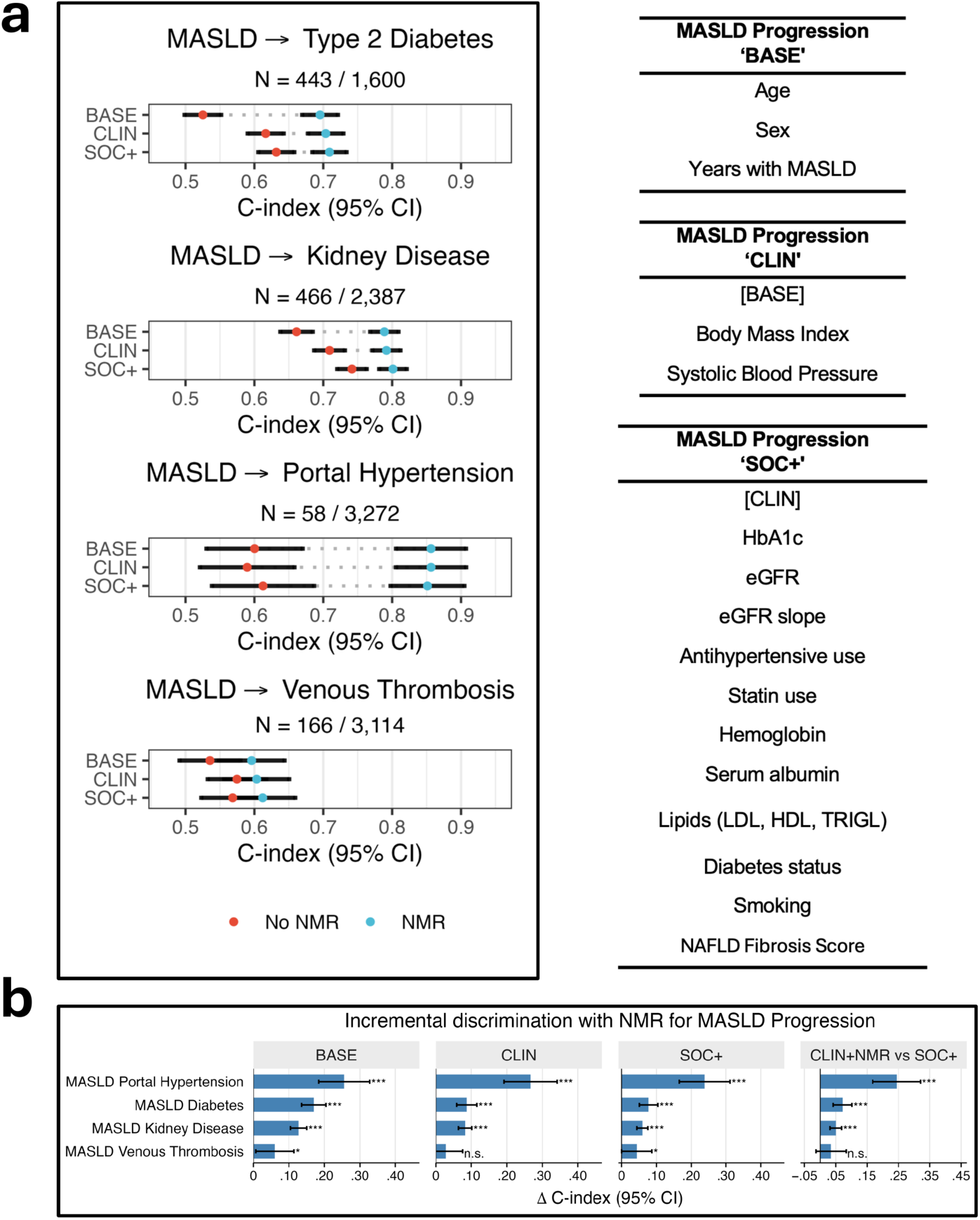
NMR metabolomics improves prediction of progression in prevalent MASLD. NMR metabolomics improves prediction of secondary outcomes in MASLD. **a**, C-index estimates and 95% confidence intervals for prediction of incident type 2 diabetes, kidney disease, portal hypertension and venous thrombosis among participants with prevalent MASLD. BASE included age, sex and MASLD duration; CLIN added BMI and systolic blood pressure; SOC+ added disease-relevant laboratory, treatment and longitudinal features. NMR-enhanced models added the 250-biomarker NMR panel to each tier. **b**, Corresponding ΔC-index estimates for addition of NMR and for CLIN+NMR versus SOC+. NMR improved discrimination across MASLD secondary outcomes, with the largest gain for portal hypertension, although this estimate was based on relatively few events.

