## Supplementary Figures for "NMR metabolomics for cardiovascular–kidney–metabolic risk stratification in an integrated healthcare system"

### Supplementary Fig. 1: Needle-to-freezer effects and correction of KPRB NMR biomarkers

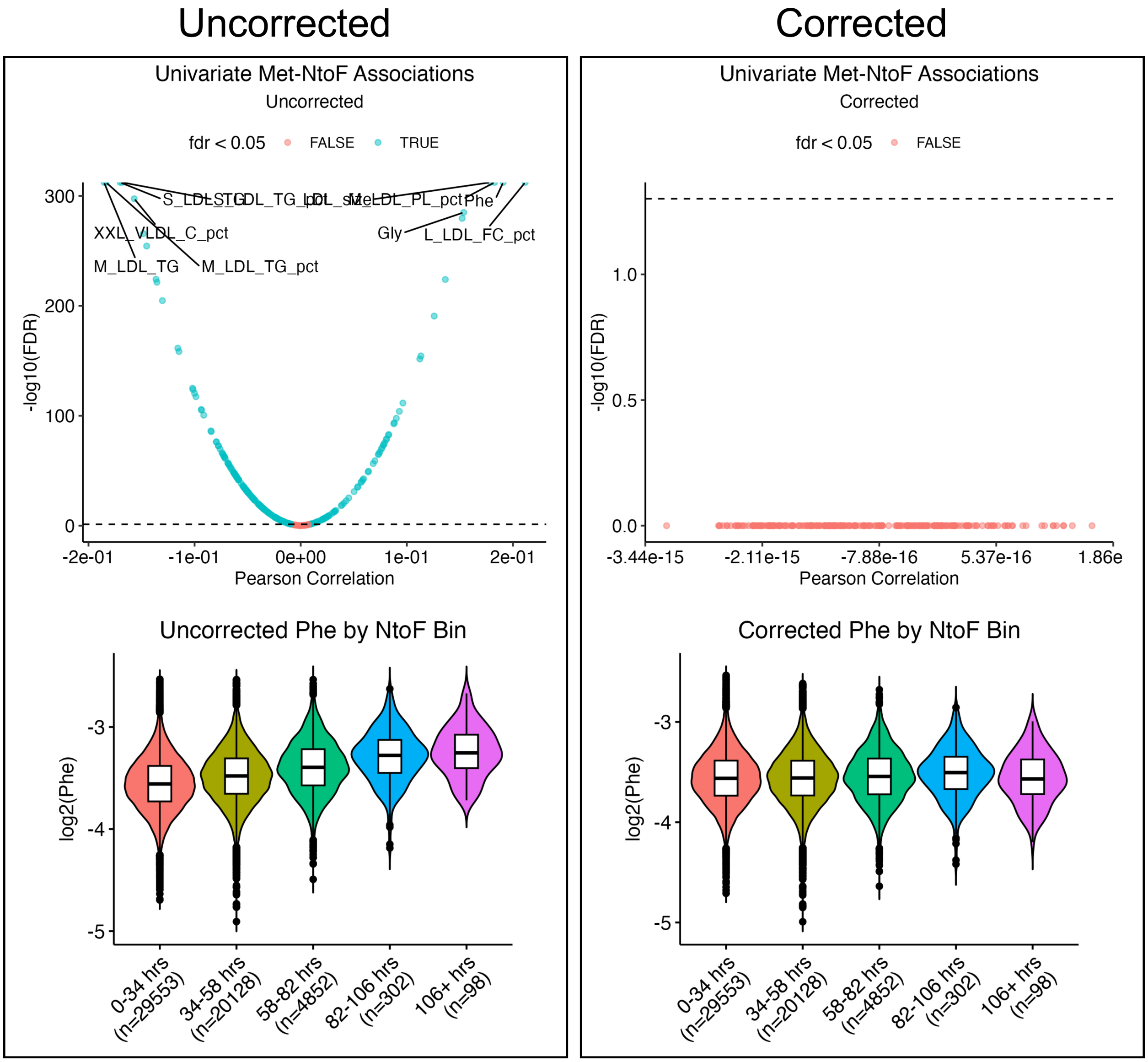


**Supplementary Fig. 1:** Pearson correlations between needle-to-freezer time and NMR metabolites before and after correction. The figure shows the metabolite-wide associations and phenylalanine as an example. Associations were generally small and were removed by residualization while preserving biomarker scale.

### Supplementary Fig. 2: Correlation structure of NMR metabolomics panel

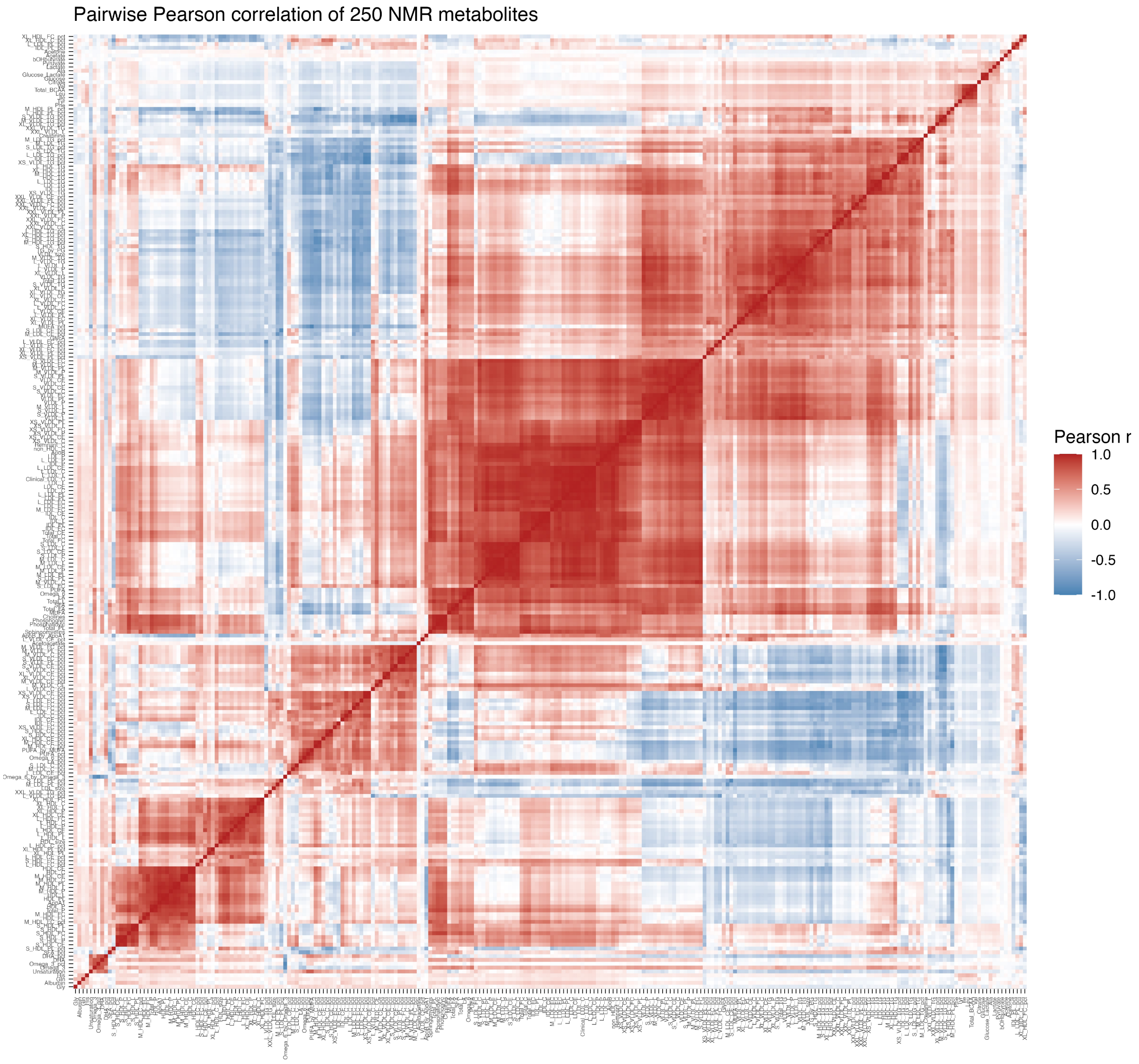


**Supplementary Fig. 2:** Pairwise correlation structure among the 250 NMR biomarkers, demonstrating strong correlation particularly within lipoprotein subclasses.

### Supplementary Fig. 3: NMR-enhanced COPD risk prediction within and across cohorts

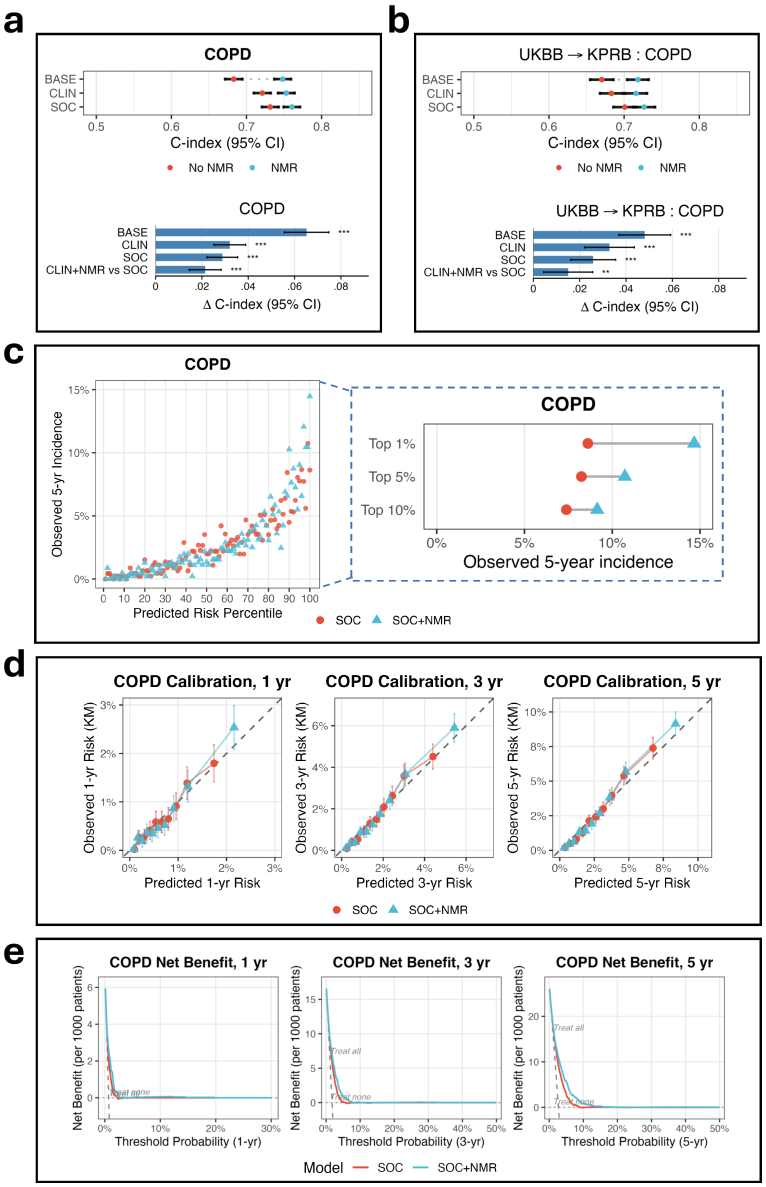


**Supplementary Fig. 3:** NMR-enhanced incident COPD prediction in the KPRB. **a**, Incident COPD risk prediction in KPRB. Top, C-index estimates for BASE, CLIN, and SOC (CLIN plus smoking status and LDL cholesterol) models, with and without the NMR panel. Bottom, corresponding C-index differences, including CLIN+NMR versus SOC. **b**, C-index differences for models trained in UK Biobank and evaluated in KPRB. **c**, High-risk enrichment for SOC and SOC+NMR models. **d**, Calibration and **e**, decision-curve analysis of SOC and SOC+NMR models at 1, 3 and 5 years. Error bars indicate 95% confidence intervals. P values were obtained from paired C-index comparisons and adjusted using the Benjamini–Hochberg procedure. P<0.05, P<0.01, P<0.001 for FDR-corrected P values.

#### Supplementary Fig. 4: NMR prediction gains are largely maintained when removing samples with missing clinical data

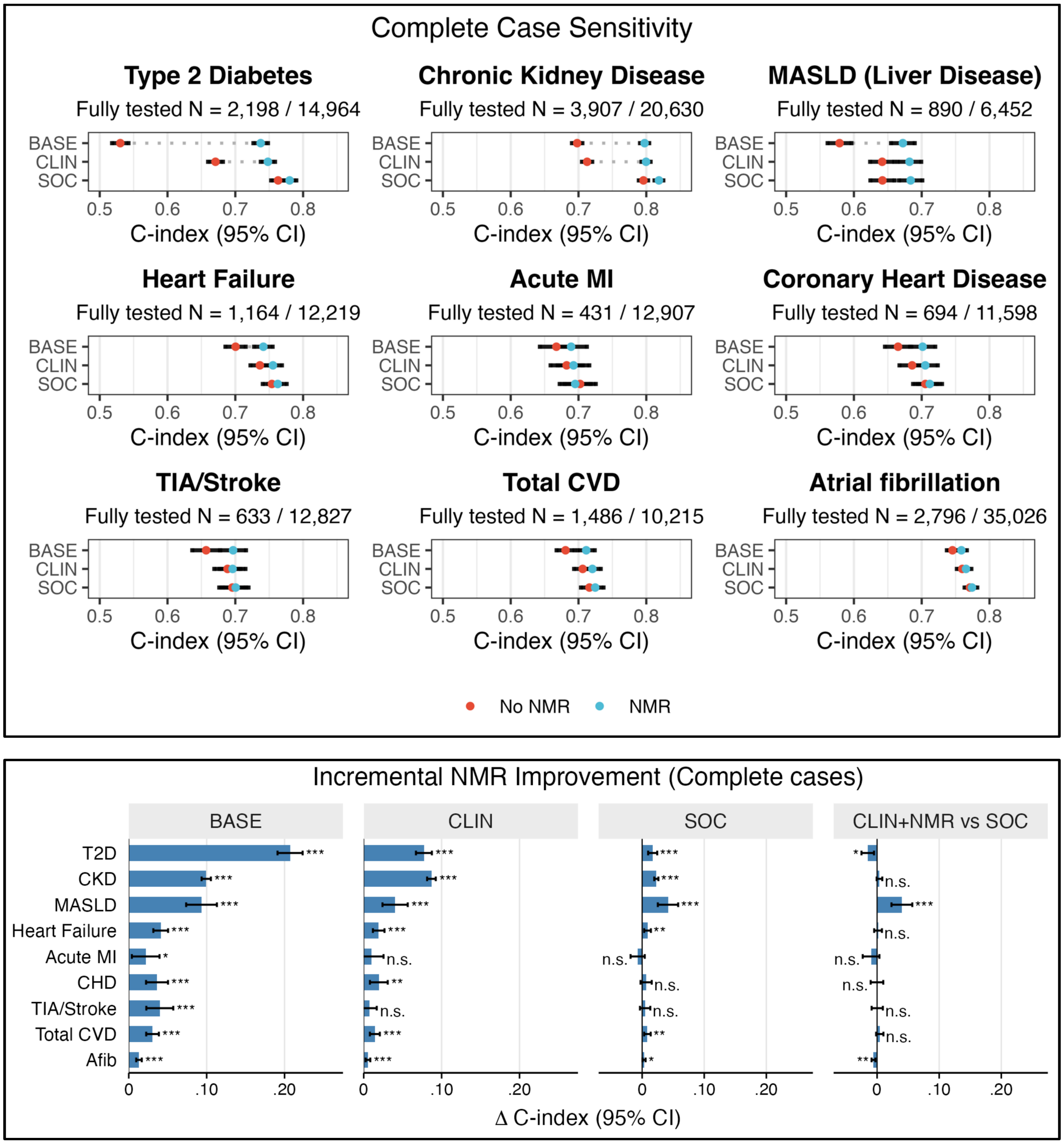


**Supplementary Fig. 4:** Complete-case sensitivity analysis of NMR-enhanced risk prediction. C-index estimates and incremental NMR improvement among participants with complete measurements across the full SOC predictor set. NMR-associated gains were broadly preserved for T2D, CKD and MASLD and remained significant for heart failure, Total CVD, and Afib. Estimates were attenuated for Acute MI, CHD and TIA/Stroke, for which complete-case restriction most substantially reduced the available event counts.

### Supplementary Fig. 5: Sensitivity analysis excluding early incident diagnoses

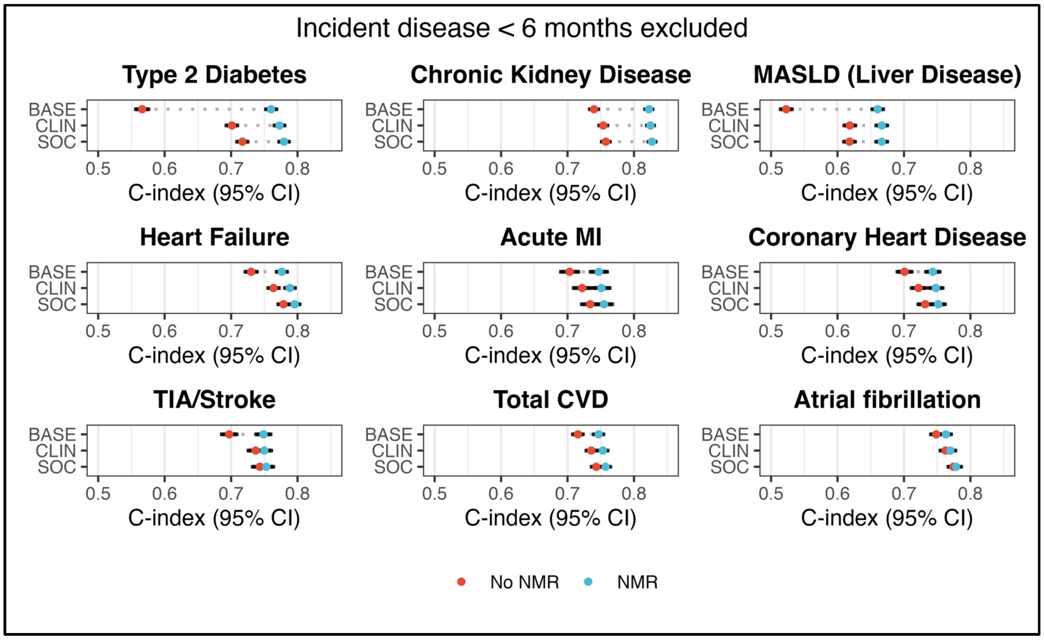


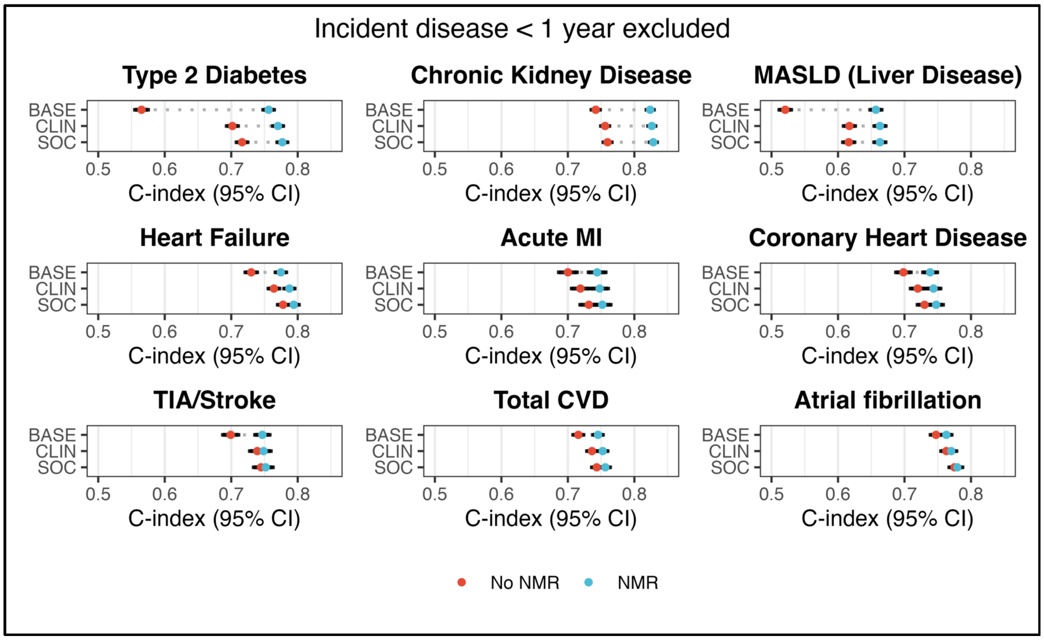


**Supplementary Fig. 5:** Model discrimination after excluding events occurring within 6 months (top panel) or 1 year (bottom panel) of sampling. Absolute discrimination decreased modestly compared to the main analysis, while the incremental improvement from NMR remained similar.

### Supplementary Fig. 6: Sensitivity analysis restricted to Kaiser participants actively engaged in healthcare system

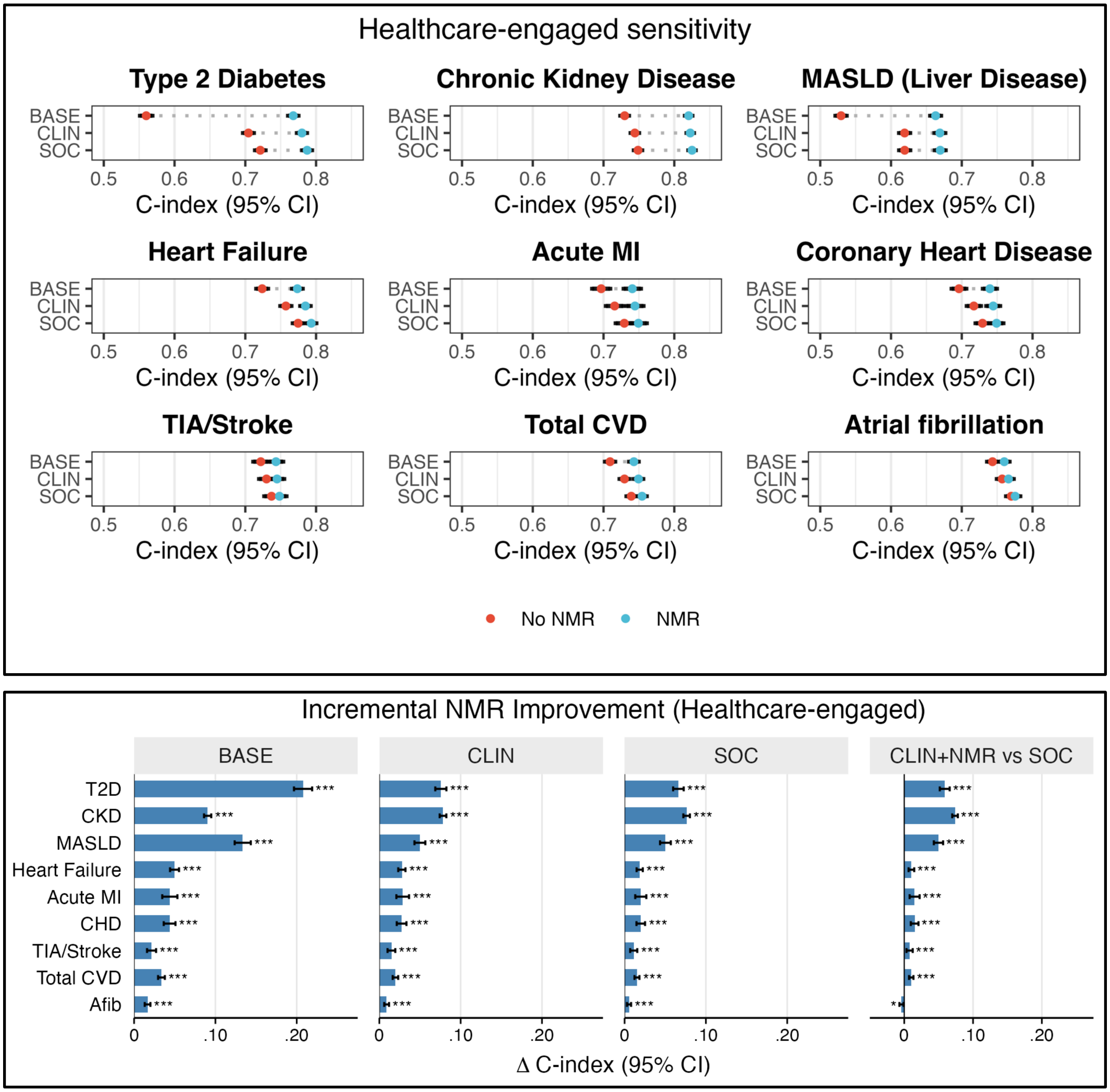


**Supplementary Fig. 6:** Model performance after excluding participants without healthcare encounters during the 2 years before sampling. NMR-associated improvements in discrimination were maintained compared to the main analysis

### Supplementary Fig. 7: Observed 5-year disease incidence across predicted risk percentiles

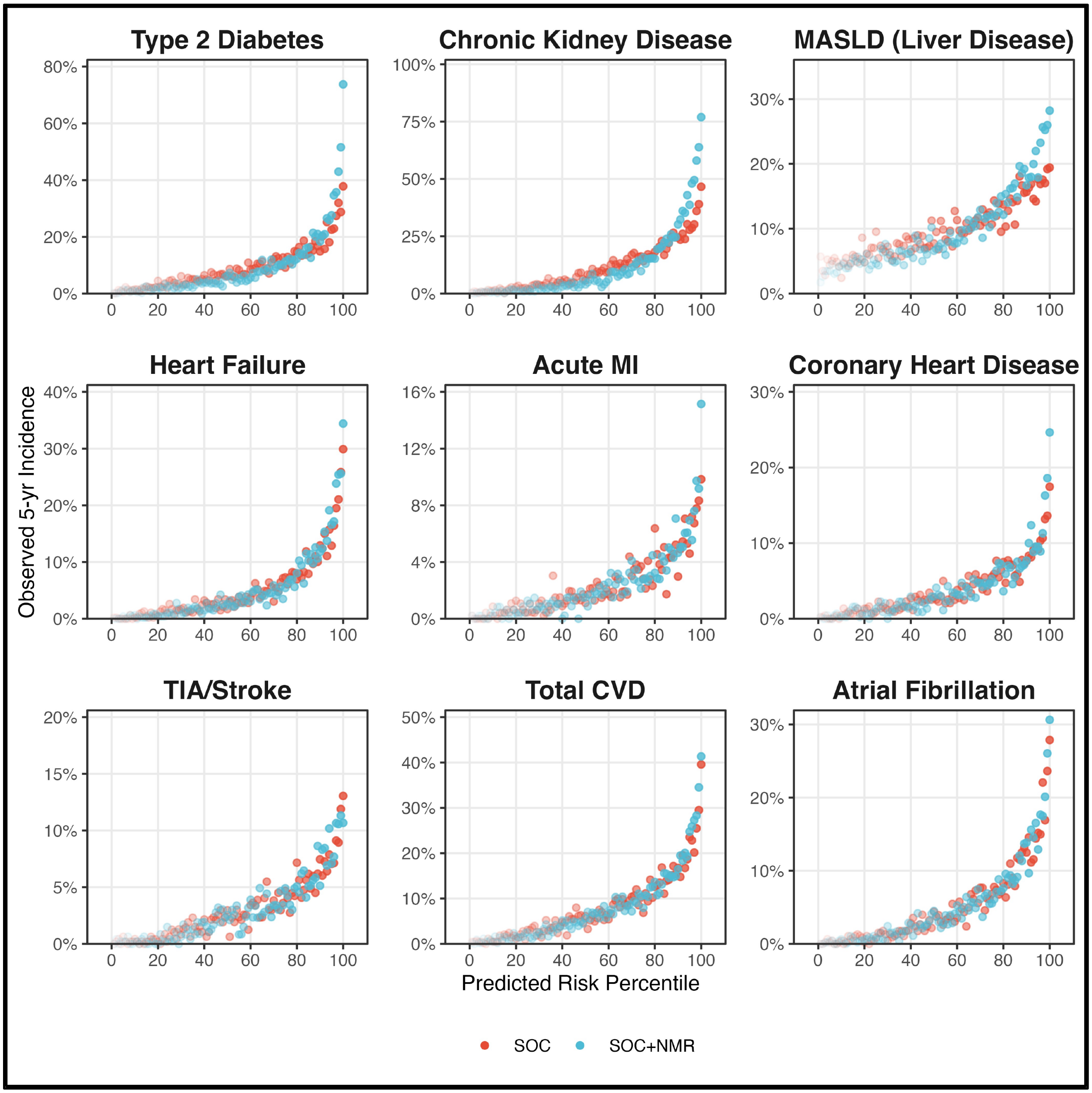


**Supplementary Fig. 7:** Observed 5-year incidence across predicted-risk percentiles for nine cardiovascular-kidney-metabolic syndrome endpoints. SOC+NMR generally produces greater enrichment of incident cases at the highest predicted risks than SOC alone.

### Supplementary Fig. 8: Cross-cohort generalizability of UK Biobank-trained NMR risk models

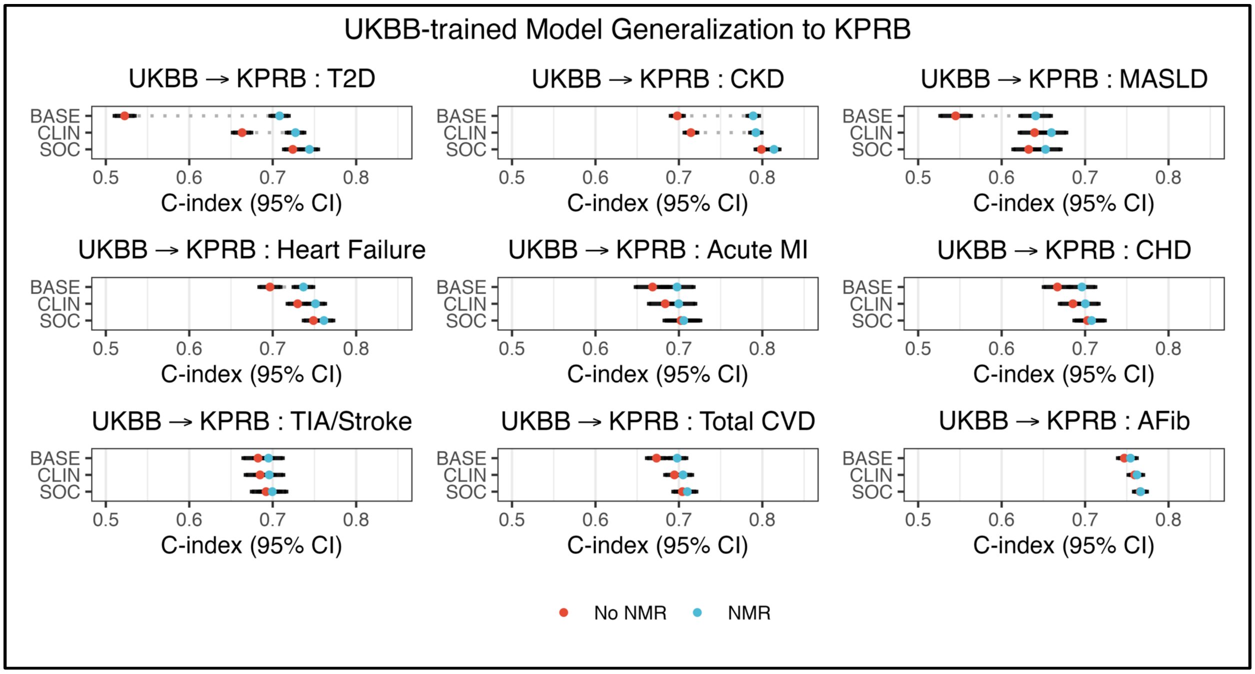


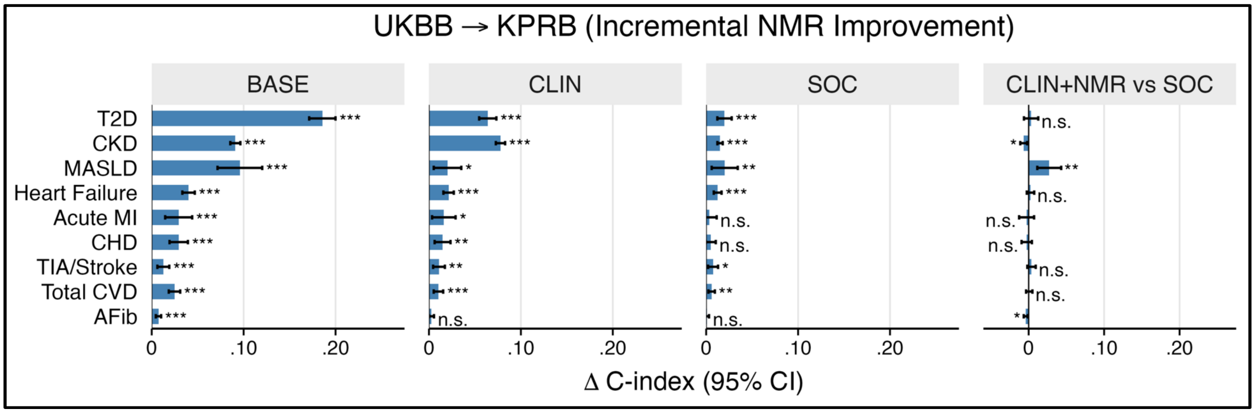


**Supplementary Fig. 8:** Cross-cohort generalizability of UK Biobank-trained NMR risk models. Top, C-index estimates and 95% confidence intervals for BASE, CLIN and SOC models, with and without NMR, trained in UK Biobank and applied without refitting to KPRB. Bottom, corresponding ΔC-index estimates for addition of NMR to each predictor tier and for CLIN+NMR versus SOC. Absolute discrimination was lower in KPRB than in within-cohort analyses, while NMR-enhanced models generally retained higher discrimination than their corresponding clinical models – with the exceptions of Acute MI, CHD, and Afib.

### Supplementary Fig. 9: NMR-enhanced models preserve calibration for secondary T2D, CKD, and MASLD outcomes

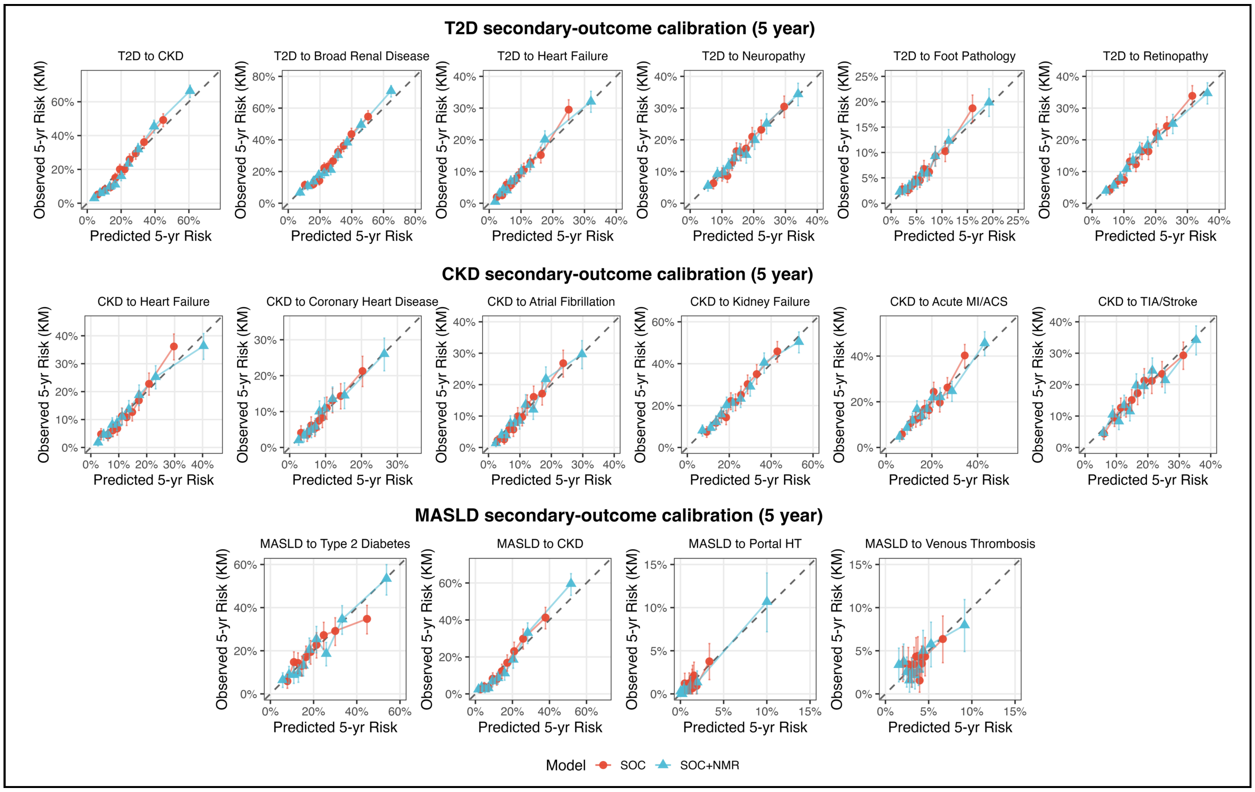


**Supplementary Fig. 9:** Predicted versus observed 5-year risk for SOC+ and NMR-enhanced SOC+ models of secondary outcomes. Top, complications among participants with prevalent T2D; middle, CKD; bottom, MASLD. Participants were grouped into deciles of predicted risk; points show mean predicted risk versus Kaplan–Meier observed incidence, with 95% confidence intervals. Dashed diagonal lines indicate perfect calibration.

### Supplementary Fig. 10: High-risk enrichment with NMR-enhanced prediction after T2D, CKD and MASLD

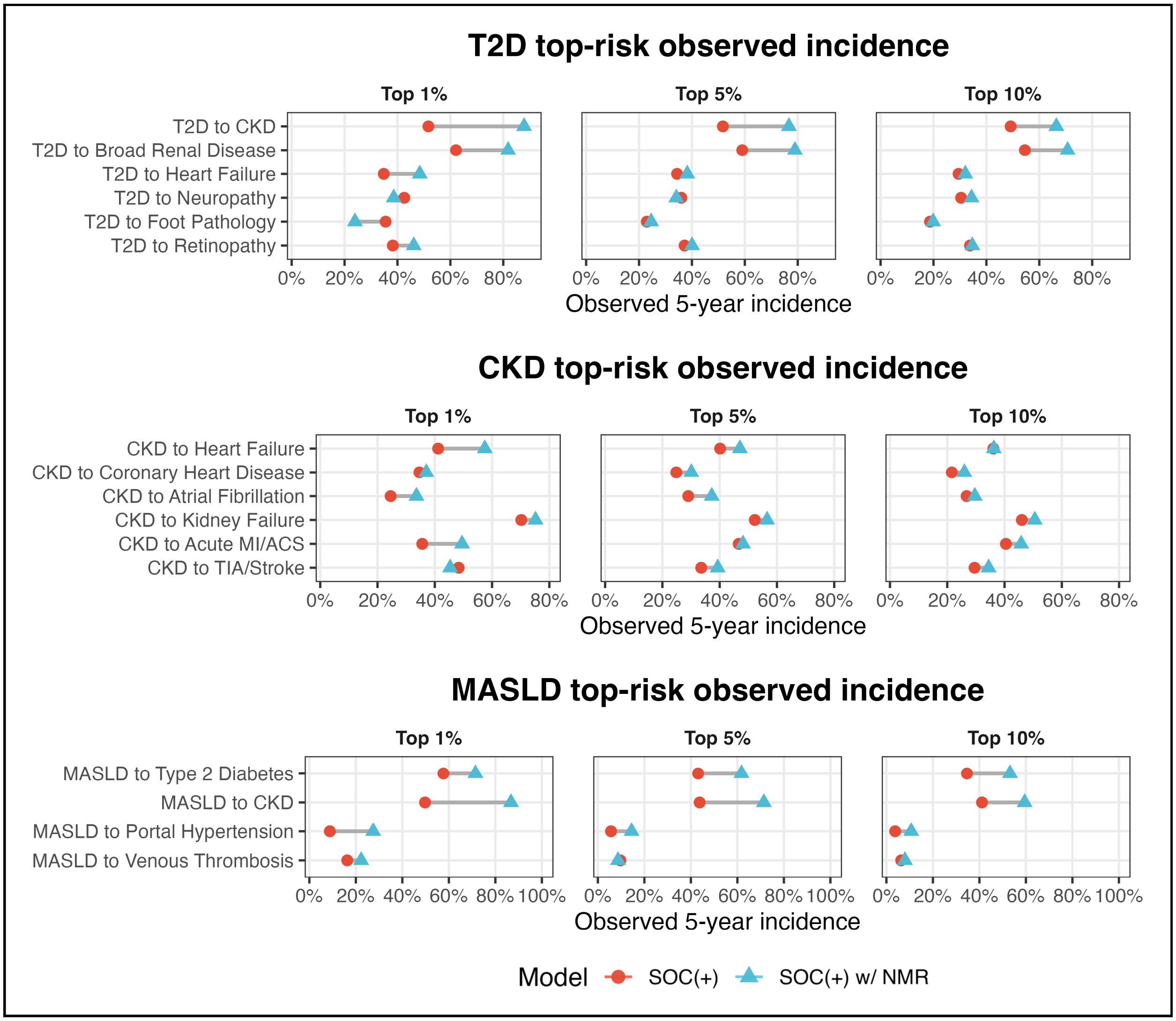


**Supplementary Fig. 10:** Observed 5-year incidence across top 1%, 5%, and 10% predicted-risk strata for SOC+ and NMR-enhanced SOC+ models. Top, complications among participants with prevalent T2D; middle, CKD; bottom, MASLD. NMR-enhanced models generally concentrated more incident events among participants at highest predicted risk, with the largest gains for renal outcomes in T2D and for kidney disease and portal hypertension in MASLD.

### Supplementary Fig. 11: Cross-cohort differences in NMR biomarker distributions

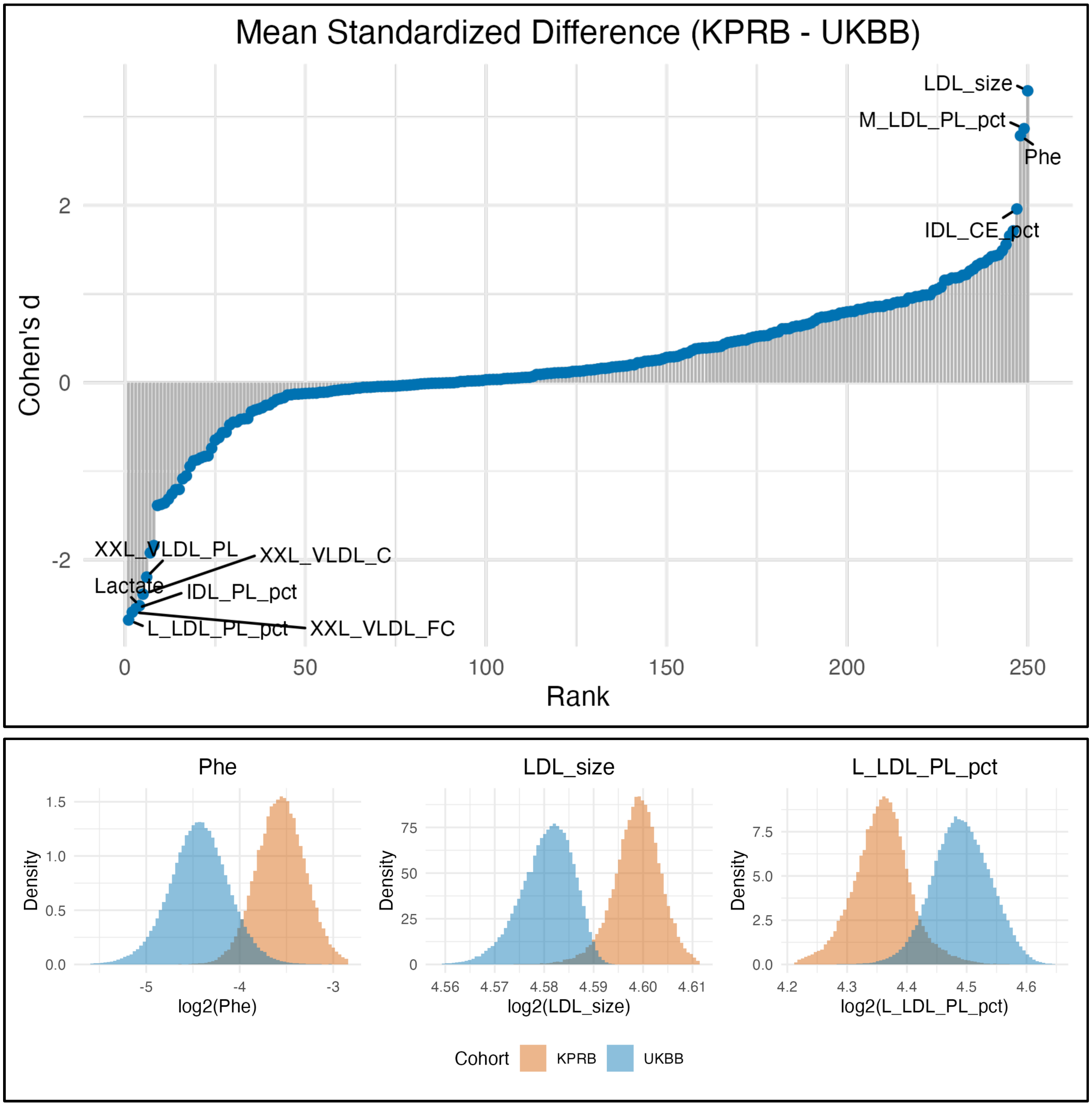


**Supplementary Fig. 11:** Standardized mean differences in NMR biomarker concentrations between KPRB serum and UK Biobank plasma quantified using Cohen's d (top panel). Representative examples of NMR biomarkers with large Cohen's d (bottom panel). For visualization purposes, the top and bottom 99.9% and 0.1% of observations were omitted in the histogram subplots.
